# The role of metabolic comorbidity in COVID-19 mortality of middle-aged adults. The case of Mexico

**DOI:** 10.1101/2020.12.15.20244160

**Authors:** Lenin Dominguez-Ramirez, Francisco Rodriguez-Perez, Francisca Sosa-Jurado, Gerardo Santos-Lopez, Paulina Cortes-Hernandez

**Author notes:** **Correspondence to:** Paulina Cortes-Hernandez M.D. Ph.D. Instituto Mexicano del Seguro Social (IMSS), Centro de Investigacion Biomedica de Oriente (CIBIOR), Cell Biology. 74360 Metepec, Puebla, Mexico. Contributed equally (Lenin Dominguez-Ramirez, Francisco Rodriguez-Perez).

## Abstract

**Background:** In contrast to developed countries, in Mexico more than half of COVID-19 deaths have occurred in adults <65-years-old, with at least a million years of life lost to premature mortality (YLL) in eight months. Mexico has a young population but a high prevalence of metabolic diseases like obesity and diabetes that contribute to COVID-19 adverse outcomes. COVID-19 could be particularly risky in population with specific comorbidity combinations that haven’t been analyzed.

**Methods:** To explore what contributes to the high COVID-19 mortality in Mexican middle-aged adults, we calculated age-stratified COVID-19 case fatality rates, YLL and relative risk (RR) of 9 comorbidities and 23 comorbidity combinations in a retrospective Mexican cohort with 905,579 PCR-confirmed COVID-19 cases/89,167 deaths, until Nov/2/2020.

**Findings:** Chronic kidney disease (CKD) had the highest RR for COVID-19 fatality, followed by diabetes and immunosuppression, that in turn had higher RR than obesity or hypertension as single comorbidities. The combination diabetes/hypertension with or without obesity had RR as high as CKD as a single comorbidity (>3 in <60-year-olds). Notably, the RR of comorbidities decreased with age, tending to values near 1 after age 60; suggesting that in Mexico, comorbidities increase COVID-19 fatality mostly in young and middle-aged adults. Our analysis suggests that advanced metabolic disease, marked by multimorbidity (more than one chronic disease per individual) or diabetes before age 60, contribute particularly to the younger age of COVID-19 fatalities in Mexico. Around 38% of YLL to COVID-19, were attributable to the synergy between COVID-19 and pre-existing diseases, mainly combinations between obesity, diabetes and hypertension. Yet, ¼ of deaths and 1/3 of YLL have occurred in individuals without known comorbidities.

**Conclusions:** The Mexican COVID-19 outbreak illustrates that middle-aged adults 45-64-yo can have high COVID-19 mortality during large outbreaks, especially if they present chronic metabolic comorbidities, but also in their absence, making them an important group of concern after elders. COVID-19 mortality in middle-aged adults is likely proportional to the gradual decline in health that accompanies ageing, which presents earlier in poorer populations that also get more exposed to SARS-CoV-2 and have less access to specialized medical attention.

## Introduction

Since the initial SARS-CoV-2 outbreaks, advanced age, male gender and chronic disease have been associated with adverse outcomes [1-6], but less is known about COVID-19 fatalities in young populations [7]. Mexico, with a population younger than world-average (median age 29.3 years-old, yo), is undergoing a large COVID-19 outbreak with high mortality, similar to that of the USA, Italy and the UK, despite having only around 1/3 of the proportion of >65-yo of these countries [9-10].

SARS-CoV-2 infections were first detected in Mexico in February 2020 and the first COVID-19 death on March/18, 2020 [11]. Eight months later, despite mitigation strategies, Mexico is the fourth country in cumulative COVID-19 deaths nearing 100,000; it is eight in mortality with almost 80 deaths/100,000 inhabitants [10] and COVID-19 ranks among the three leading causes of death in the country, along with cardiovascular disease and diabetes [12]. Additionally, Mexico has the second highest COVID-19 case-fatality in the world (after Yemen) [11], and more than half of the fatalities have been in adults younger than 65-yo [7].

To explore what contributes to the high COVID-19 fatality in middle-aged adults in Mexico, we analyzed the first eight months of the outbreak and its relation to preexisting chronic disease. In Mexico, 36% of adults are obese, 39% overweight, 10-14% diabetic, while 22% have diagnosed hypertension [13-14]. Thus, the Mexican COVID-19 outbreak includes a large share of young and middle-aged adults, with a variety of comorbidity combinations. Our analysis is illustrative of the role of chronic comorbidities in COVID-19 fatality at different ages and of the impact of COVID-19 in young populations.

## Methods

We analyzed COVID-19 datasets updated by Mexican health authorities, available at [15], that include all symptomatic patients evaluated as probable COVID-19, since outbreak start, along with their comorbidities and outcomes (retrospective cohort). Datasets arise from a national epidemiological surveillance system with standardized reporting, described in [16], under World Health Organization (WHO) case definitions. Patient identity was absent from datasets and information was managed adhering to the ethical principles of the Helsinki declaration. We analyzed datasets at regular intervals, from the first available through Nov/2/2020 (end of study), when 905,579 cases had been confirmed as SARS-CoV-2 by RT-qPCR; of which 89,167 (9.8%) had died (Study diagram in Figure 1). Cases determined as COVID-19 through epidemiological/clinical evidence, without RT-PCR were not included.

**Figure 1.**
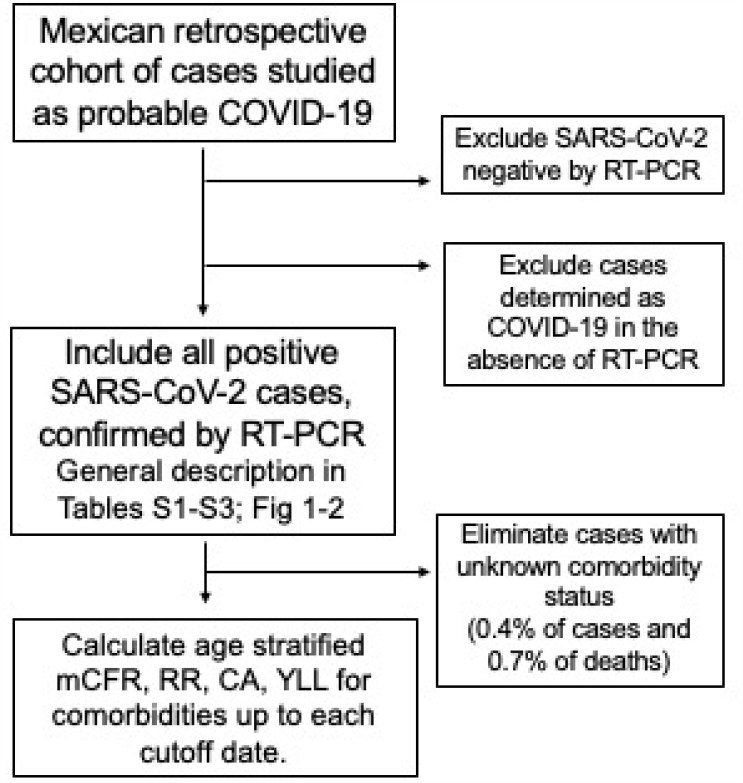
Study Design. mCFR, measured case fatality rate; RRd relative risk of COVID-19 death; CA correspondence analysis; YLL, years of life lost due to premature (COVID-19) mortality. Confirmed cases/deaths included at each cutoff date with known comorbidity status: Aug/6/2020 (first cutoff) 50,182/460,803; Sept/12/2020 (second cutoff) 70,139/661,254; Nov/2/2020 (end of sudy) 88,610/901,664.

Measured Case fatality rates (mCFR) were calculated as the percent of cumulative COVID-19 cases that died, by date reported; and mortality as the cumulative number of COVID-19 deaths in 100,000 inhabitants. Relative Risk of death (RRd) and 95% CI (confidence interval) were calculated in GraphPad Prism version 7.0a, with Koopman asymptotic score, excluding cases and deaths with unknown comorbidity status, at cutoff dates Aug/6 and Sept/12, 2020, that marked >50,000 and >70,000 COVID-19 deaths in Mexico and comprise the first COVID-19 peak in the country (Fig 2A). Demographic and medical characteristics of COVID-19 cases and deaths at cutoff dates and at the end of study (Nov/2/2020), are in Tables S1-S3. Few differences in mCFR and RR were found between cutoff dates. RR of comorbidities in figures and tables are for the second cutoff (Sept/12/2020), except when indicated. Student’s T and Fisher’s exact tests were used for numerical and categorical variables, respectively. Years of life lost to premature mortality (YLL) were determined for each individual under the age of 70 that died of COVID-19, with the formula:

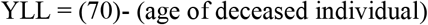

**Figure 2.**
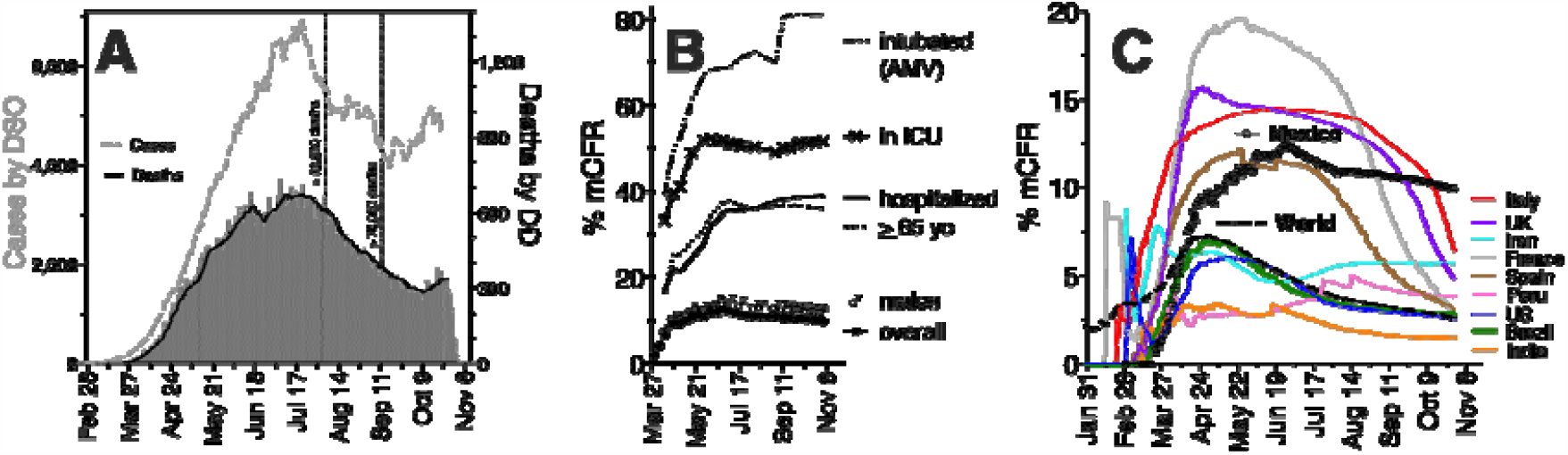
Eight-month evolution of the SARS-CoV-2 outbreak in Mexico (Feb/27-Nov/2, 2020). **(A)** COVID-19 cases and deaths confirmed by RT-PCR until Nov/2/2020. Daily deaths are in bars and weekly rolling averages in lines; for cases, by day-of-symptom-onset (DSO) and for deaths, by day-of-death (DD), excluding the last 10 days that could be artificially low due to information delay. Dotted vertical lines mark cutoff dates used in subsequent analyses: Aug/6/2020 and Sept/12/2020. **(B)** Evolution of measured Case Fatality Rate (mCFR); AMV, assisted mechanical ventilation; ICU, intensive care unit). **(C)** mCFR in Mexico vs the top ten countries with the most cumulative COVID-19 deaths, calculated from cumulative cases and deaths by date reported.

Multivariate correspondence analyses (CA) of categorical variables and Hierarchical Clustering of Principal Components (HCPC) with the k-means method, were performed in R through Rstudio, with libraries factoextra and FactoMineR [17], as in [18].

## Results

### Eight-month evolution of the COVID-19 outbreak in Mexico (Feb/27 to Nov/2, 2020)

COVID-19 incidence first grew in Mexico from March to July/2020, descended in Aug-Sept and a potential second peak is evolving since Oct/2020 (Figure 2A). In parallel, the measured case fatality rate (mCFR), increased sharply during March-April and more slowly during May-June, peaking at 12.4% in late-June (Figure 2B-C), with a slight descent since (9.9% by Nov/2/2020). PCR-positivity has fluctuated above 40% since April (not shown), suggesting a high rate of undetected cases that contributes to the large mCFR.

In Mexico, the vast majority (>97%) of COVID-19 cases have been treated in public health services (not shown). Hospitalized patients have a high mCFR (>35% since July, Figure 2B) and a high Relative Risk of death (RRd >21 *vs* the non-hospitalized), that increases with worsening clinical conditions (Figure 2B; Tables S1-S3). The majority of deaths (55.3%) have been in individuals hospitalized in non-critical areas, while 11% occurred out-of-hospital (Table S3). Of hospitalized cases, 20% received critical care (assisted mechanical ventilation, and/or treatment in an intensive care unit) and account for 33.3% of total deaths (Table S3). The fatality rate in critical areas has been >50% in ICU (Intensive Care Unit) and 70-80% for patients under AMV (assisted mechanical ventilation) since July (Figure 2B). Mexico’s mCFR has not decreased after peak incidence as much as in other countries (Figure 2C), likely driven by low case detection, high hospital fatality [19] and differences in reporting; for example, only symptomatic COVID-19 cases are included in the Mexican datasets.

### COVID-19 demography during the first wave, in Mexico

More COVID-19 deaths have happened in males (64%, Tables S1-S3), who have higher mCFR (Figure 3B) and a RRd around 1.65 *vs* females, that peaks at ages 20-59 (Figure 4A-B). COVID-19 mortality, mCFR and RRd increase with age (Figure 3 and 4A) and deceased individuals were older in average than cases (Figure 3A, Tables S1-S4). Yet, in contrast to high income countries, in Mexico more than half of COVID-19 deaths have been in adults 20-64 yo (52.7%), while adults >65-yo and children 0-19 yo account for 46.9% and 0.4% of deaths, respectively (until Nov/2/2020) (Figure 3A). Additionally, in Mexico mCFR and mortality increase at earlier ages than in high income countries, even in individuals without comorbidity (Figure 3B-E). CFR comparisons are complicated by differences in testing levels and case reporting [5,20], whereas mortality is a less ambiguous end-point adjusted by population size. Mexico’s age-stratified COVID-19 mortality was several-fold higher in 45-75-yo than in high income countries with similar large outbreaks like Italy, the USA and the UK (all four countries had similar mortality 62-80 deaths/100,000 inhabitants by Nov/2/2020 [21-23]).

**Figure 3.**
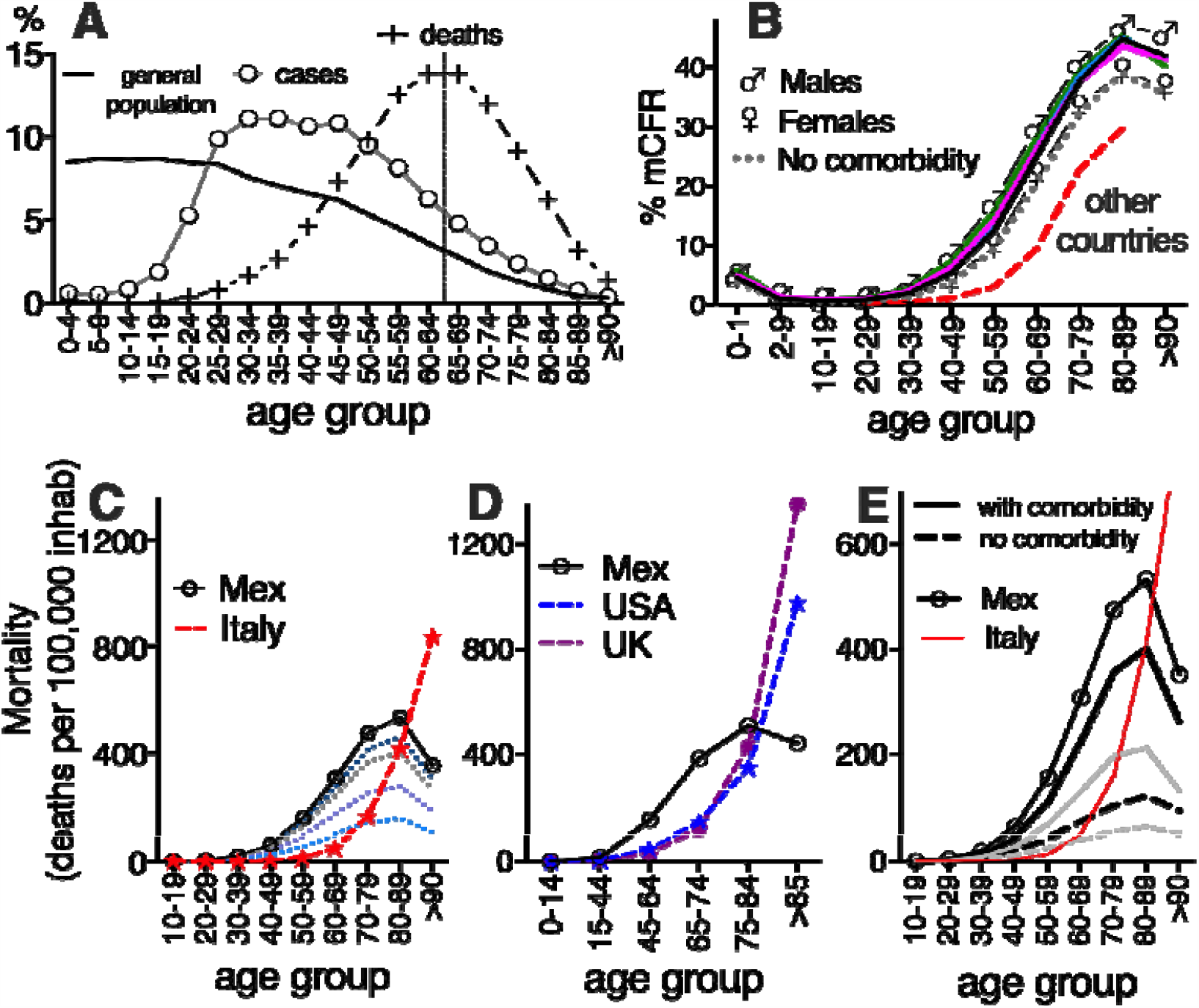
Age distribution of COVID-19 in Mexico. **(A)** Percent of COVID-19 cases and deaths per age group until Nov/2/2020 (end of study), compared to the Mexican population pyramid (2020 estimate [8]). **(B)** Measured Case Fatality Rate (mCFR) per age group, at four points in the Mexican 2020 outbreak, July/4 (green), Aug/6 (blue), Sept/12 (pink) and Nov/2 (black), with 30,000, 50,000, 70,000 and 89,000 COVID-19 deaths confirmed as SARS-CoV-2 positive by RT-PCR. The four curves overlap. At each date, overall mCFR (all ages) was 12.2%, 10.9%, 10.9.% and 9.9% respectively. Fatality rates for males, females and individuals without comorbidity are shown for Aug/6/2020 at 50,000 deaths. Broken red line represents the mCFR reported in [5], with data from China, Spain, Italy, New York and UK, during initial large outbreaks (74,000 pooled deaths and an overall mCFR of 12.1%; similar to Mexico’s). **(C-E)** COVID-19 mortality per 100,000 inhabitants of each age group, in Mexico (up to Nov/2/2020 89,000 deaths) *vs* other countries (Italy at 37,457 deaths [21]; USA at 212,328 deaths [22]; UK at 55,311 deaths [23]). In **(C)**, dotted lines mark the mortality in Mexico at different time-points in the outbreak, from bottom to top: July/4 at 30,000 deaths; Aug/6 at 50,000 deaths; Sept/12 at 70,000 deaths; and Oct/4 at 79,000 deaths, to show outbreak progression. Different age groups were used in **(C)** and **(D-E)** to adapt to the age-partitions reported by other countries. In **(E)** COVID-19 deaths in Mexico with-(solid line) and without-comorbidity (discontinuous line), were separated and mortality was calculated over the total inhabitants in each age group, at end of study: 89,000 deaths (black lines) and at 50,000 deaths (gray lines). Overall mortality at end of study is shown in circles for Mexico and in a red line for Italy, and correspond to the same curves as in **(C)**, as reference.

**Figure 4.**
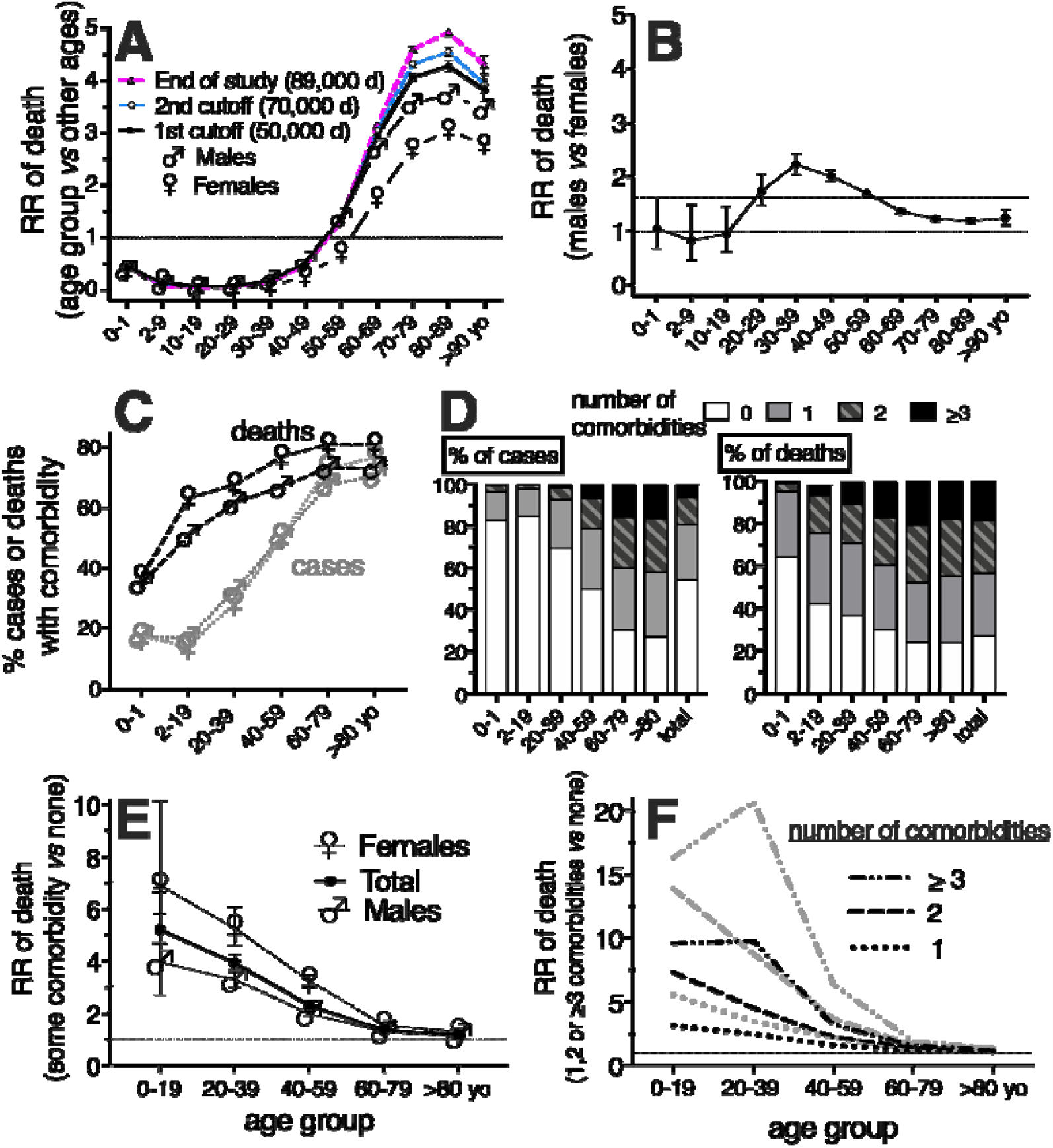
RR of COVID-19 fatality, contributed by age (A), gender (B), or chronic comorbidity (E-F) in Mexico. **(C)** Percent of COVID-19 cases (gray) and deaths (black) with at least one comorbidity. **(D)** Age-group distribution of cases and deaths, with 0, 1, 2, or >3 comorbidities. In **(F)**, gray are females and black, males. 95% CI (confidence intervals) are displayed in **(A**,**B**,**D)** as whiskers. Dotted horizontal line marks RRd=1 and in **(B)**, discontinuous horizontal line marks the overall RRd=1.62 calculated for males *vs* females (all ages). Data correspond to the cutoff date, Aug/6/2020, with >50,000 deaths; except in **(A)** where data for the second cutoff (Sept/12/2020) and for end of study (Nov/2/2020) are also shown. In **(C-F)** only individuals with known comorbidity status were included.

In Mexico, only a small proportion of the detected cases correspond to children (3.8% cases in <20-yo, Figure 3A), perhaps in relation to the suspension of in-person education since March, 2020 and to the mildness of cases in that age group, many of which may be undetected. At the other end of the spectrum, COVID-19 mortality doesn’t increase further after 85-yo (Figure 3C-D), similar to reported for India [24]. To analyze if this could be due to undercounting of COVID-19 deaths among elders (for example by incorrect assignment of death causes), we looked at excess mortality: 51,826 and 59,410 excess deaths were reported until epidemiological week 31 in 45-64 and >65-yo in Mexico [25], corresponding to 203.7 and 604.9 deaths/100,000 inhabitants in each age group, which describes a similar trend to that of confirmed COVID-19 mortality. Thus, excess mortality doesn’t suggest that COVID-19 deaths in elders are more undercounted that at other ages. However, further age-stratification of excess mortality after 65-yo would be necessary to confirm this.

The higher mCFR and mortality in middle aged-adults in Mexico *vs* high income countries could be related to behavioral, environmental, biological/health factors or to access to specialized medical attention. Since Mexico has a high prevalence of chronic metabolic disease in adults, in the next section, we explored the relation of COVID-19 fatalities to pre-existing chronic diseases (comorbidities).

### Risk of COVID-19 death contributed by pre-existing medical conditions (comorbidity) in Mexico

About 74% of individuals that died of COVID-19 had at least one preexisting chronic disease (Figure 4D; Tables S1-S3), the most common being hypertension, diabetes and obesity, in 43%, 38% and 25% deaths (and 20%, 16%, 19% cases) (Figure 5). Hypertension and diabetes were found at a similar prevalence in COVID-19 cases as in the general population [13-14]; while obesity was found at about half the prevalence in COVID-19 (Figure 5).

**Figure 5.**
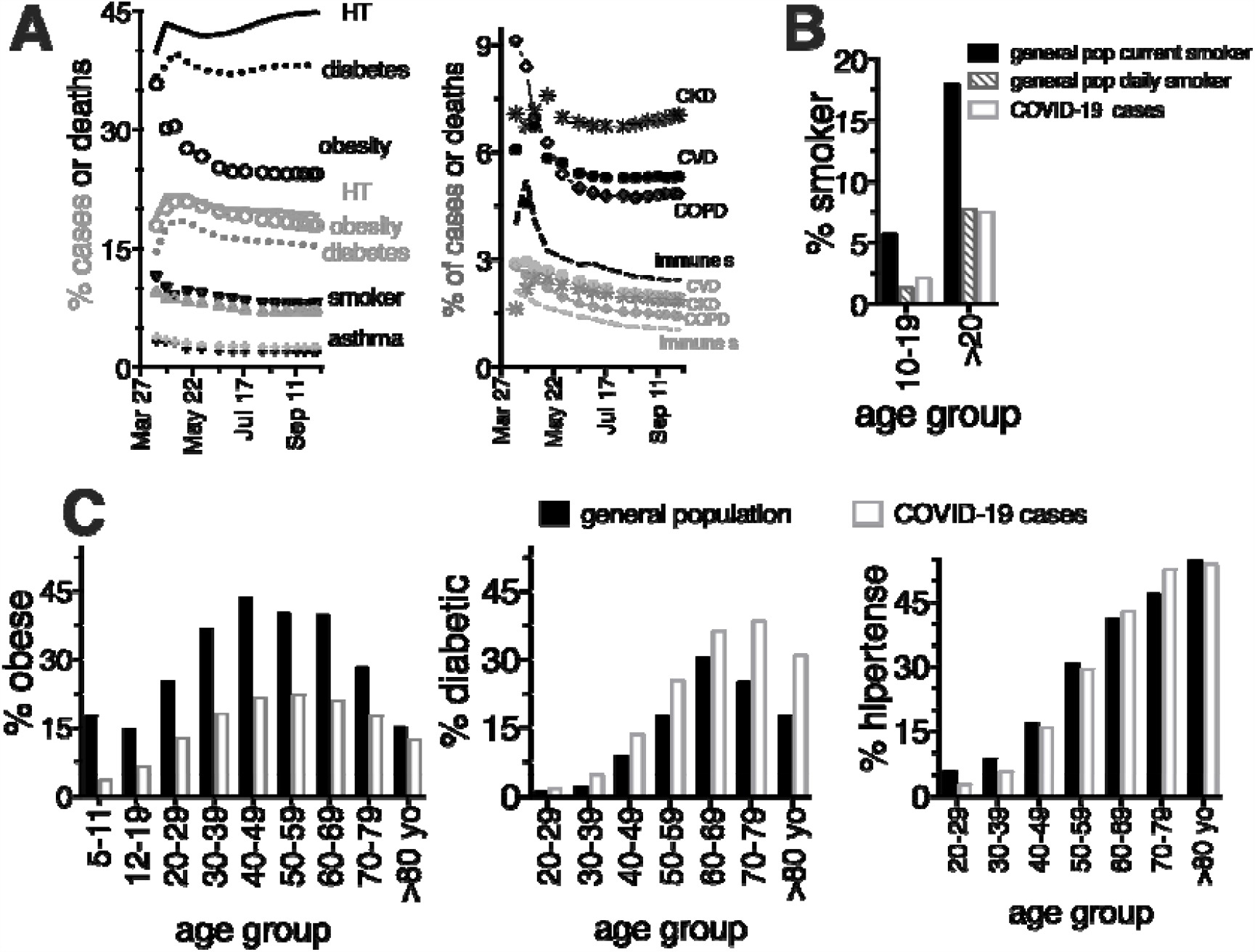
Frequency of comorbidities in COVID-19 in Mexico, **(A)** comorbidities in COVID-19 cases (gray) and deaths (black), along the 2020 Mexican outbreak. Abbreviations as in text. **(B-C)** Frequency of comorbidities in COVID-19 cases (white bars) compared with their estimated prevalence in the Mexican general population (black bars), as reported by health authorities for 2018 [13-14]. In (B), different bars show current smoker and daily smokers for the general population as reported in [14].

COVID-19 patients with comorbidity were older than patients without (Table S4) and there have been more COVID-19 deaths with comorbidity at older ages (Figure 4C-D), paralleling the increase in chronic diseases with age in the general population (Figure 5) [13-14]. The relative risk of COVID-19 death (RRd) contributed by having any comorbidity in Mexicans, at different ages is in Figure 4E-F. The most relevant finding was that the RRd of pre-existing disease decreased sharply with age, tending to values near 1 in adults >60-yo, as comorbidities were present at similar proportions in COVID-19 cases and deaths in older age groups (Figure 4C-D).

Next we analyzed the age-stratified mCFR and RRd of 9 specific comorbidities: obesity; diabetes (DM) without type distinction (>90% diabetes in Mexican adults is type II); hypertension (HT); other cardiovascular diseases (CVD); chronic kidney disease (CKD); smoking; asthma; chronic obstructive pulmonary disease (COPD); immunosuppression (ImmunoS); and unspecified “other” conditions; across genders (Table S5) and in age groups (Table 1), as single or multiple comorbidity to COVID-19. Little variation was found between cutoff dates (Table 1 vs Table S6) suggesting sufficient sampling since the first cutoff, except at pediatric ages where dispersion persists.

**Table 1.** Measured case fatality rate (mCFR) and relative risk of death (RR_d_) of chronic comorbidities by age group, in COVID-19 in Mexico (Feb/27-Sep/12, 2020; at >70,000 deaths).

| Age group/<br>disease |  |  | As a single comorbidity |  | In combination with 1 other<br>comorbidity |  | In combination with ≥2 other<br>comorbidities |  |
| --- | --- | --- | --- | --- | --- | --- | --- | --- |
|  | Cases | Deaths | %m |  | %m |  | %m |  |
|  | n (%) | n (%) | CFR | RRd [95% CI]<sup>1</sup> | CFR | RRd [95% CI]<sup>1</sup> | CFR | RRd [95% CI]<sup>1</sup> |
| 0-19 yo |  |  |  |  |  |  |  |  |
| Obesity | 1,292 (34.8) | 29 (21.2) | 1.8 | 2.7 [1.68-4.42]*** | 3.3 | 5.2 [2.5-10.7]*** | 6.5 | 10.0 [3.9-24.3]*** |
| Diabetes | 219 (5.9) | 18 (13.1) | 5.7 | 8.9 [4.3-18.0]**** | 10.0 | 15.5 [7.1-31.9]**** | 13.5 | 20.9 [9.0-44.3]**** |
| HT | 181 (4.9) | 14 (10.2) | 4.5 | 7.0 [2.4-19.7]** | 8.1 | 12.5 [5.3-27.7]**** | 11.3 | 17.5 [8.1-35.7]**** |
| CVD | 209 (5.6) | 11 (8.0) | 4.0 | 6.2 [2.6-14.3]** | 4.6 | 7.2 [2.4-20.0]** | 15.0 | 23.3 [8.0-57.0]*** |
| CKD | 129 (3.5) | 15 (10.9) | 5.7 | 8.9 [3.4-21.7]** | 22.9 | 35.4 [18.3-62.3]**** | 12.5 | 19.4 [6.7-48.9]*** |
| Smoker | 469 (12.6) | 7 (5.1) | 0.8 | 1.3 [0.43-3.72]<sup>ns</sup> | 2.6 | 4.0 [1.10-14.1]<sup>ns</sup> | 8.0 | 12.4 [3.4-39.2]* |
| Asthma | 891 (24.0) | 3 (2.2) | 0.3 | 0.4 [0.11-1.53]<sup>ns</sup> | 0.8 | 1.24 [0.22-6.8]<sup>ns</sup> | - | - |
| COPD | 0 (0.0) | 0 (0.0) | - | - | - | - | - | - |
| ImmunoS | 411 (11.1) | 31 (22.6) | 4.9 | 7.5 [4.24-13.2]**** | 11.6 | 17.9 [10.6-29.5]**** | 11.6 | 18.0 [7.7-38.7]**** |
| Other | 601 (16.2) | 63 (46.0) | 10.5 | 16.2 [11.6-22.6]**** | 10.3 | 15.9 [9.9-25.1]**** | 11.4 | 17.6 [7.6-37.9]**** |
| w/comorb | 3,713 (100) | 137 (100) | 3.0 | 4.6 [3.6-6.0]**** | 6.9 | 10.6 [7.3-15.3]**** | 11.3 | 17.6 [9.8-30.5]**** |
| 20-39 yo |  |  |  |  |  |  |  |  |
| Obesity | 39,176 (52.4) | 1,336 (54.0) | 2.6 | 3.2 [2.92-3.49]**** | 3.8 | 4.6 [4.1-5.1]**** | 9.5 | 11.5 [10.2-13.1]**** |
| Diabetes | 8,372 (11.2) | 705 (28.5) | 5.7 | 7.0 [6.05-8.04]**** | 8.2 | 10.0 [8.8-11.4]**** | 13.8 | 16.8 [14.9-19.0]**** |
| HT | 11,756 (15.7) | 799 (32.3) | 2.6 | 3.2 [2.67-3.86]**** | 7.2 | 8.8 [7.8-9.8]**** | 13.5 | 16.4 [14.7-18.3]**** |
| CVD | 1,466 (2.0) | 99 (4.0) | 3.1 | 3.8 [2.39-5.92]**** | 4.5 | 5.5 [3.6-8.4]**** | 13.8 | 16.8 [13.2-21.2]**** |
| CKD | 2,212 (3.0) | 399 (16.1) | 9.2 | 11.2 [8.86-14.1]**** | 18.9 | 23.0 [19.8-26.7]**** | 26.9 | 32.8 [28.6-37.6]**** |
| Smoker | 20,929 (28.0) | 296 (12.0) | 0.5 | 0.63 [0.50-0.80]**** | 2.3 | 2.8 [2.4-3.4]**** | 6.6 | 8.1 [6.6-9.9]**** |
| Asthma | 7,129 (9.5) | 124 (5.0) | 0.9 | 1.04 [0.76-1.43]<sup>ns</sup> | 2.4 | 2.9 [2.1-3.8]**** | 5.5 | 6.7 [4.9-9.1]**** |
| COPD | 520 (0.7) | 40 (1.6) | 3.7 | 4.5 [2.07-9.52]** | 4.2 | 5.1 [2.5-10.2]*** | 14.3 | 17.4 [12.2-24.5]**** |
| ImmunoS | 1,518 (2.0) | 167 (6.8) | 6.0 | 7.4 [5.36-10.1]**** | 9.9 | 12.1 [9.1-16.0]**** | 18.6 | 22.6 [18.5-27.5]**** |
| Other | 4,060 (5.4) | 230 (9.3) | 3.2 | 3.9 [3.11-4.91]**** | 6.8 | 8.3 [6.6-10.3]**** | 13.1 | 16.0 [12.9-19.7]**** |
| w/comorb | 74,759 (100) | 2,474 (100) | 2.3 | 2.8 [2.62-3.04]**** | 5.1 | 6.2 [5.7-6.8]**** | 12.0 | 14.6 [13.2-16.2]**** |
| 40-59 yo |  |  |  |  |  |  |  |  |
| Obesity | 56,562 (43.6) | 7,283 (42.9) | 9.4 | 1.7 [1.63-1.78]**** | 12.5 | 2.3 [2.2-2.4]**** | 20.3 | 3.7 [3.5-3.8]**** |
| Diabetes | 48,487 (37.4) | 8,884 (52.3) | 14.0 | 2.5 [2.44-2.66]**** | 17.7 | 3.2 [3.1-3.3]**** | 24.2 | 4.4 [4.2-4.6]**** |
| HT | 56,776 (43.8) | 8,835 (52.1) | 8.9 | 1.6 [1.54-1.70]**** | 15.7 | 2.8 [2.7-3.0]**** | 23.5 | 4.3 [4.1-4.4]**** |
| CVD | 4,150 (3.2) | 732 (4.3) | 7.8 | 1.4 [1.11-1.80]** | 12.8 | 2.3 [2.0-2.7]**** | 23.5 | 4.3 [3.9-4.6]**** |
| CKD | 4,657 (3.6) | 1,686 (9.9) | 20.4 | 3.7 [3.17-4.32]**** | 31.0 | 5.6 [5.1-6.2]**** | 41.5 | 7.5 [7.2-7.9]**** |
| Smoker | 17,208 (13.3) | 1,777 (10.5) | 5.2 | 0.9 [0.85-1.04]<sup>ns</sup> | 11.1 | 2.0 [1.87-2.2]**** | 19.0 | 3.4 [3.2-3.7]**** |
| Asthma | 6,702 (5.2) | 556 (3.3) | 3.9 | 0.7 [0.59-0.86]*** | 7.5 | 1.4 [1.18-1.6]**** | 15.0 | 2.7 [2.4-3.0]**** |
| COPD | 2,509 (1.9) | 552 (3.3) | 15.5 | 2.8 [2.30-3.41]**** | 16.8 | 3.1 [2.6-3.6]**** | 27.9 | 5.1 [4.6-5.6]**** |
| ImmunoS | 2,973 (2.3) | 601 (3.5) | 12.9 | 2.3 [1.96-2.78]**** | 16.8 | 3.1 [2.6-3.5]**** | 27.5 | 5.0 [4.5-5.5]**** |
| Other | 6,413 (4.9) | 1,099 (6.5) | 11.3 | 2.1 [1.83-2.29]**** | 15.6 | 2.8 [2.6-3.2]**** | 25.5 | 4.6 [4.3-5.0]**** |
| w/comorb | 129,627 (100) | 16,973 (100) | 9.9 | 1.8 [1.74-1.85]**** | 15.0 | 2.7 [2.6-2.8]**** | 23.0 | 4.2 [4.0-4.3]**** |
| 60-79 yo |  |  |  |  |  |  |  |  |
| Obesity | 22,071 (28.2) | 7,678 (29.3) | 29.1 | 1.22 [1.16-1.28]**** | 33.0 | 1.38 [1.33-1.43]**** | 38.6 | 1.61 [1.56-1.66]**** |
| Diabetes | 41,240 (52.7) | 14,861 (56.7) | 30.7 | 1.28 [1.24-1.33]**** | 34.8 | 1.45 [1.41-1.49]**** | 40.9 | 1.71 [1.66-1.75]**** |
| HT | 51,588 (66.0) | 17,629 (67.3) | 27.5 | 1.15 [1.11-1.19]**** | 34.0 | 1.42 [1.38-1.46]**** | 40.3 | 1.68 [1.64-1.73]**** |
| CVD | 5,800 (7.4) | 2,169 (8.3) | 27.6 | 1.15 [1.01-1.31]* | 33.1 | 1.38 [1.28-1.49]**** | 40.5 | 1.69 [1.62-1.77]**** |
| CKD | 4,837 (6.2) | 2,341 (8.9) | 39.9 | 1.67 [1.43-1.91]**** | 45.6 | 1.91 [1.77-2.05]**** | 49.8 | 2.1 [2.00-2.16]**** |
| Smoker | 7,947 (10.2) | 2,877 (11.0) | 29.3 | 1.22 [1.14-1.31]**** | 33.4 | 1.40 [1.31-1.48]**** | 41.6 | 1.74 [1.66-1.81]**** |
| Asthma | 2,353 (3.0) | 633 (2.4) | 22.4 | 0.94 [0.79-1.10]<sup>ns</sup> | 25.1 | 1.05 [0.91-1.20]<sup>ns</sup> | 29.7 | 1.24 [1.14-1.36]**** |
| COPD | 4,985 (6.4) | 1,978 (7.6) | 32.4 | 1.35 [1.21-1.50]**** | 36.8 | 1.54 [1.43-1.65]**** | 42.9 | 1.79 [1.71-1.88]**** |
| ImmunoS | 2,087 (2.7) | 789 (3.0) | 27.5 | 1.15 [0.96-1.35]<sup>ns</sup> | 34.5 | 1.44 [1.29-1.61]**** | 42.7 | 1.78 [1.66-1.91]**** |
| Other | 4,127 (5.3) | 1,747 (6.7) | 37.8 | 1.58 [1.45-1.72]**** | 41.6 | 1.74 [1.62-1.86]**** | 44.8 | 1.87 [1.78-1.97]**** |
| w/comorb | 78,222 (100) | 26,190 (100) | 29.1 | 1.22 [1.19-1.25]**** | 34.4 | 1.44 [1.40-1.47]**** | 40.3 | 1.68 [1.64-1.73]**** |
| ≥80 yo |  |  |  |  |  |  |  |  |
| Obesity | 2,080 (16.6) | 960 (16.5) | 43.5 | 1.12 [0.97-1.27]<sup>ns</sup> | 45.6 | 1.17 [1.06-1.28]** | 47.2 | 1.21 [1.13-1.30]**** |
| Diabetes | 5,267 (42.1) | 2,499 (43.0) | 44.7 | 1.15 [1.05-1.24]** | 47.1 | 1.21 [1.14-1.28]**** | 49.1 | 1.26 [1.19-1.33]**** |
| HT | 9,177 (73.4) | 4,270 (73.5) | 44.5 | 1.14 [1.08-1.20]**** | 46.2 | 1.18 [1.13-1.25]**** | 49.5 | 1.27 [1.20-1.34]**** |
| CVD | 1,666 (13.3) | 759 (13.1) | 44.6 | 1.14 [0.96-1.33]<sup>ns</sup> | 43.3 | 1.11 [0.99-1.23]<sup>ns</sup> | 46.9 | 1.20 [1.11-1.30]**** |
| CKD | 826 (6.6) | 443 (7.6) | 49.2 | 1.26 [0.96-1.57]<sup>ns</sup> | 52.2 | 1.34 [1.16-1.52]*** | 54.7 | 1.40 [1.29-1.52]**** |
| Smoker | 1,384 (11.1) | 658 (11.3) | 42.3 | 1.08 [0.94-1.24]<sup>ns</sup> | 50.5 | 1.30 [1.16-1.43]**** | 48.0 | 1.23 [1.13-1.34]**** |
| Asthma | 322 (2.6) | 97 (1.7) | 23.1 | 0.59 [0.35-0.93]* | 29.2 | 0.75 [0.53-1.01]<sup>ns</sup> | 32.6 | 0.84 [0.67-1.02]<sup>ns</sup> |
| COPD | 1,769 (14.1) | 831 (14.3) | 46.0 | 1.18 [1.03-1.33]* | 49.2 | 1.26 [1.15-1.38]**** | 45.9 | 1.18 [1.09-1.27]*** |
| ImmunoS | 323 (2.6) | 158 (2.7) | 46.6 | 1.19 [0.88-1.52]<sup>ns</sup> | 44.3 | 1.14 [0.87-1.42]<sup>ns</sup> | 51.6 | 1.32 [1.14-1.51]*** |
| Other | 911 (7.3) | 511 (8.8) | 59.8 | 1.53 [1.34-1.72]**** | 58.2 | 1.49 [1.33-1.65]**** | 53.4 | 1.37 [1.25-1.50]**** |
| w/comorb | 12,508 (100) | 5,812 (100) | 44.8 | 1.15 [1.10-1.20]**** | 46.9 | 1.20 [1.15-1.26]**** | 48.9 | 1.25 [1.19-1.32]**** |

<sup>1</sup> Some patients had more than one comorbidity; thus the sum of comorbidities exceeds the number of individuals with comorbidity (w/comorb). Disease abbreviations as in text.
<sup>2</sup> RR were calculated against individuals with no comorbidities in the same age group. A total of 661,255 cases and 70,139 COVID-19 deaths with known comorbidity status were included. \*\*\*\* p<0.0001; \*\*\* p<0.001; \*\* p<0.01; \* p<0.05; ns, not significant

Comorbidity in COVID-19 deaths was more frequent in females (79% vs 70% in males; age distribution in Figure 4C) and females had on average more comorbidities per individual than males (2.03±0.53 in females and 1.88±1.0 in males, p<0.0001, in patients that died from COVID-19). Females had a slightly higher frequency of all comorbidities, except for smoking, which was 2-3 times more frequent in males (Table S5). The RRd of comorbidity was larger in females (Figure 4E-F; Table S5). In both genders the risk was higher with more comorbidities per individual but tended to 1 in older groups (Figure 4F).

At early ages, “other” diseases, obesity, immunosuppression and asthma predominated. Smoking and obesity rose in young adults, particularly as single comorbidity; whereas diabetes, hypertension, COPD and CVD increased at later ages (Figure 6A). The proportion of COVID-19 deaths with one comorbidity remained stable across ages at ∼30%, but comorbidity combinations increased with age (Figure 4D). In particular, patients with ≥3 comorbidities were infrequent at pediatric ages but represented 16-20% of COVID-19 cases and deaths in ≥40-yo (Figure 4D). The frequency of specific combinations of 2 or ≥3 chronic comorbidities in COVID-19 deaths per age group is displayed in Figure 6B, Figures S1-S2 and Table S7. The small group of pediatric patients that died from COVID-19 and had more than one comorbidity, presented combinations mostly between immune disease, “other” diseases and/or CKD (Figures S1-S2). In contrast, in adults the most common comorbidity combinations were between obesity, diabetes and/or hypertension, sometimes with smoking, CKD, CVD or COPD, in accord to their prevalence in the general population.

**Figure 6.**
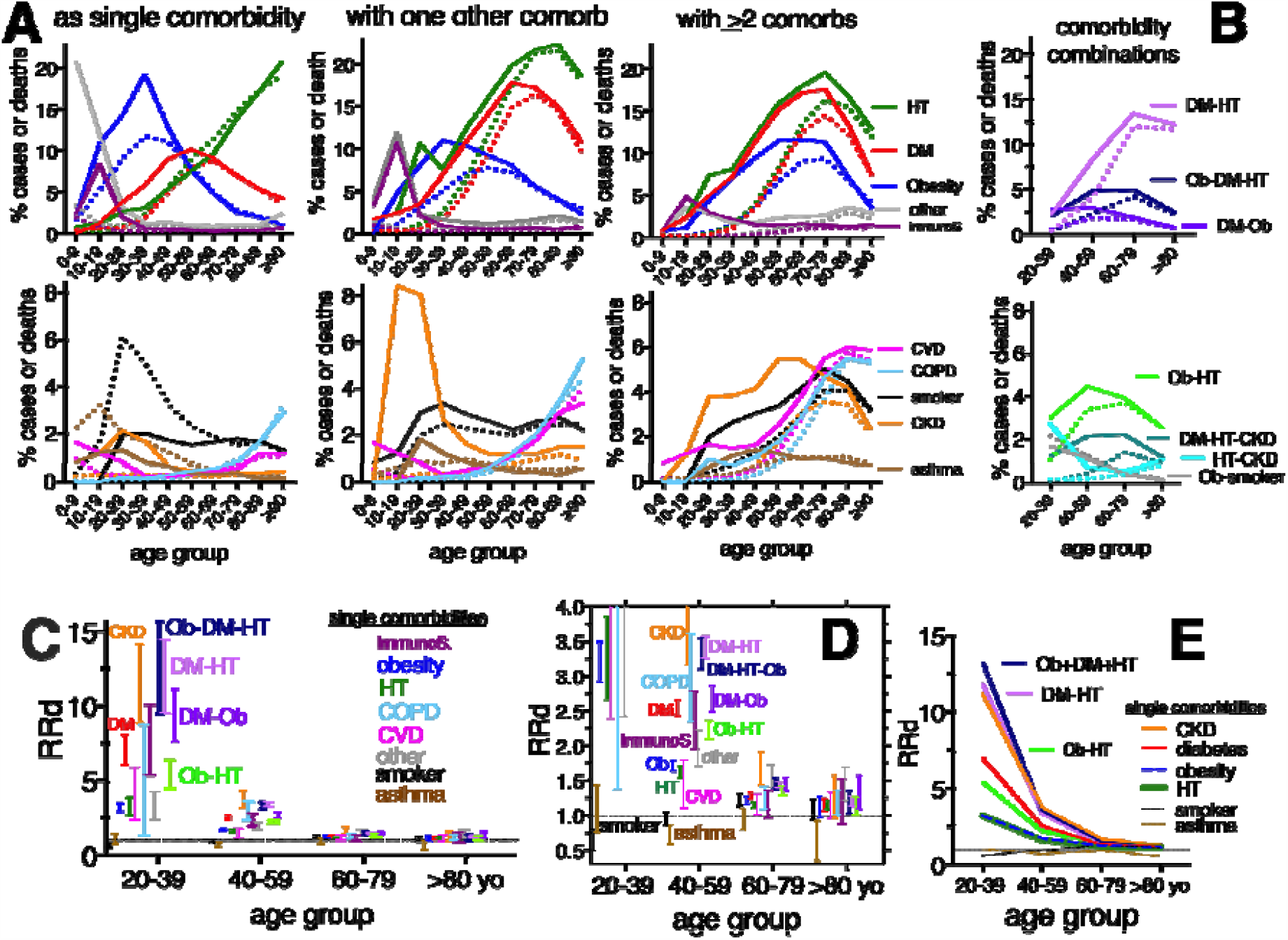
Frequency (A-B) and RRd (C-E) of specific comorbidities for COVID-19 in Mexico across age groups. In **(A-B)**, COVID-19 cases (solid line) and deaths (dotted line), with each comorbidity, either as single or as multimorbidity, are displayed preserving the color code for each disease. **(B)** shows the most frequent disease combinations found as COVID-19 comorbidity. **(C-E)** are different visualizations of the RRd found: **(C-D)** display the 95% CI, and (**E**) the trend across ages. Single comorbidities are compared to the common comorbidity combinations: obesity/diabetes/hypertension (Ob-DM-HT); diabetes/hypertension (DM-HT); diabetes/obesity (DM-Ob) and obesity/hypertension (Ob-HT). The first two combinations overlap with the RRd of CKD as a single comorbidity (orange). **(D)** Amplifies a fraction of **(C)** for improved visualization of RRd in age groups >40-yo. Horizontal dotted line marks RRd=1. Data correspond to the second cutoff date Sept/12/2020 (>70,000 deaths), excluding individuals with unknown comorbidity status. Disease abbreviations as in text.

Age-stratified RRd from specific comorbidities and their combinations is illustrated in Figure 6C-E, and detailed in Tables 1-2, always using as reference individuals in the same age group without comorbidity. The RRd increased with multi-morbidity and decreased steeply with age, tending to values near 1 in older adults, even with multiple diseases. CKD, as single or multiple comorbidity, had the largest mCFRs and RRds for COVID-19 death at all ages, most prominent in young- and middle-aged adults (Tables 1-2, Figure 6C-E). Next came immunosuppression and diabetes, that as single comorbidities, at least doubled the risk of death from COVID-19 in <60-yo, (95% CI >5 in 20-39 yo and >2 in 40-59 yo). Obesity and hypertension as single comorbidities had lower RRd than diabetes, still substantial in young adults (95% CI, 2.7-3.9 in 20-39 yo and 1.5-1.8 in 40-59 yo) (Table 1). In children 5-19 yo, 5.7% of COVID-19 cases and 15.7% of deaths had obesity, peaking at ages 12-19 mostly as a single comorbidity (80% cases and 61% deaths with obesity in children had only that comorbidity). The RRd from obesity as a single comorbidity in children was 2.7 (95% CI, 1.7-4.4), despite a low mCFR of 1.8%, and increased with more comorbidities (Table 1).

In adults, comorbidity combinations that included diabetes tended to have larger RRd than combinations with hypertension or obesity (Tables 1-2, Figure 6C-E, Figure S1). The most common comorbidity combination in COVID-19 cases and deaths in Mexico was diabetes/hypertension (DM/HT), that increased with age (peak at 60-79 yo in 13.4% of deaths and 12% of cases, Figure 6B). In turn, up to 6.5% of COVID-19 cases and 8% of deaths had diabetes/hypertension/obesity, sometimes with smoking and/or CKD (Figure 4b; Figure S2). The prevalence of this triad in COVID-19 cases and deaths, peaked at ages 50-79 (Figure 6C; Figure S2). The RRd of combinations of diabetes/hypertension with or without obesity was as high as that of CKD as a single comorbidity (RRd>3 in <60-yo; Figure 6C-E; Table 2). Next in frequency came the combination Obesity/Hypertension (Ob/HT) that had lower RRd than diabetes as a single or multiple comorbidity (Figure 6B-D, Tables 1-2). Obesity’s prevalence in the general population peaks in 40-49 yo and then decreases. Accordingly, COVID-19 comorbidity combinations that included obesity, also peaked at younger ages than other combinations.

**Table 2.** Measured case fatality rate (mCFR) and relative risk of COVID-19 death (RR_d_) of the 23 most common comorbidity combinations by age group, in Mexico (Feb/27-Sept/12, 2020; second cutoff at >70,000 deaths).

| Comorbidity | 20-39 yo |  | 40-59 yo |  | 60-79 yo |  | ≥80 yo |  |
| --- | --- | --- | --- | --- | --- | --- | --- | --- |
|  | %m |  | %m |  | %m |  | %m |  |
|  | CFR | RR<sub>d</sub> [95% CI]<sup>1</sup> | CFR | RR<sub>d</sub> [95% CI]<sup>1</sup> | CFR | RR<sub>d</sub> [95% CI]<sup>1</sup> | CFR | RR<sub>d</sub> [95% CI]<sup>1</sup> |
| No comorbidity | 0.8 | - | 5.5 | - | 23.9 | - | 39.0 | - |
| DM-HT | 9.6 | 11.8 [9.5-14.4]**** | 18.8 | 3.4 [3.25-3.57]**** | 34.3 | 1.4 [1.39-1.47]**** | 46.6 | 1.2 [1.13-1.27]**** |
| DM-obesity | 7.5 | 9.2 [7.6-11.1]**** | 14.7 | 2.7 [2.49-2.87]**** | 34.6 | 1.4 [1.35-1.54]**** | 51.8 | 1.3 [1.09-1.57]** |
| Obesity-HT | 4.4 | 5.4 [4.46-6.4]**** | 12.3 | 2.2 [2.10-2.36]**** | 32.5 | 1.4 [1.29-1.42]**** | 44.1 | 1.1 [1.01-1.26]* |
| DM-CKD | 21.4 | 26.1 [16.3-39.6]**** | 40.4 | 7.3 [6.31-8.44]**** | 52.6 | 2.2 [1.97-2.43]**** | 43.8 | 1.1 [0.72-1.56]ns |
| HT-CKD | 23.6 | 28.7 [24.0-34.0]**** | 31.5 | 5.7 [5.04-6.47]**** | 41.1 | 1.7 [1.53-1.91]**** | 55.9 | 1.4 [1.22-1.65]**** |
| Obesity-CKD | 7.1 | 8.7 [4.3-17.1]**** | 19.8 | 3.6 [2.45-5.13]**** | 39.6 | 1.7 [1.13-2.24]* | 50.0 | 1.3 [0.24-2.33]ns |
| CKD-ImmuneS | 21.4 | 26.1 [16.3-39.6]**** | 22.0 | 4.0 [2.18-6.67]*** | 28.6 | 1.2 [0.34-2.68]ns | 33.3 | 1.7 [0.53-2.42]ns |
| DM-smoker | 6.7 | 8.2 [5.2-12.7]**** | 14.7 | 2.7 [2.31-3.08]**** | 35.6 | 1.5 [1.33-1.65]**** | 59.7 | 1.5 [1.23-1.81]*** |
| HT-smoker | 4.1 | 5.0 [3.0-8.35]**** | 10.1 | 1.8 [1.53-2.20]**** | 30.9 | 1.3 [1.16-1.43]**** | 45.0 | 1.2 [0.97-1.34]ns |
| Obesity-smoker | 2.0 | 2.5 [1.98-3.1]**** | 10.2 | 1.8 [1.66-2.06]**** | 30.9 | 1.3 [1.12-1.47]*** | 60.0 | 1.5 [1.08-1.94]* |
| DM-COPD | 8.7 | 10.6 [2.94-32.7]* | 20.0 | 3.6 [2.64-4.89]**** | 40.2 | 1.7 [1.47-1.90]**** | 46.2 | 1.2 [0.93-1.45]ns |
| HT-COPD | 8.0 | 9.7 [2.7-30.46]* | 17.9 | 3.3 [2.42-4.30]**** | 38.3 | 1.6 [1.43-1.77]**** | 49.8 | 1.3 [1.12-1.44]*** |
| Obesity-COPD | 3.5 | 4.3 [1.17-14.5]ns | 16.1 | 2.9 [2.08-4.03]**** | 31.1 | 1.3 [1.02-1.61]* | 37.1 | 1.0 [0.59-1.38]ns |
| Obesity-asthma | 2.8 | 3.4 [2.36-4.7]**** | 7.4 | 1.3 [1.06-1.68]**** | 22.9 | 1.0 [0.69-1.29]ns | - | - |
| HT-CVD | 4.8 | 5.9 [2.3-14.33]** | 10.5 | 1.9 [1.47-2.48]**** | 32.1 | 1.3 [1.21-1.48]**** | 42.5 | 1.1 [0.96-1.23]ns |
| Ob-DM-HT | 10.8 | 13.2 [10.6-16.3]**** | 19.4 | 3.5 [3.34-3.73]**** | 36.0 | 1.5 [1.44-1.57]**** | 48.8 | 1.3 [1.12-1.39]*** |
| Ob-DM-HT-CKD | 28.6 | 34.8 [18.6-57.5]**** | 41.6 | 7.6 [6.57-8.58]**** | 51.5 | 2.2 [1.94-2.36]**** | 39.3 | 1.0 [0.60-1.48]ns |
| Ob-DM-HT-smk | 4.4 | 5.4 [2.1-13.13]** | 18.4 | 3.3 [2.80-3.98]**** | 43.0 | 1.8 [1.60-2.01]**** | 62.2 | 1.6 [1.18-1.96]** |
| Ob-DM-HT-CVD | 4.8 | 5.8 [1.03-27.7]ns | 25.0 | 4.5 [3.69-5.52]**** | 38.0 | 1.6 [1.41-1.77]**** | 41.3 | 1.1 [0.79-1.35]ns |
| Ob-DM-HT-COPD | 42.9 | 52.2 [19.3-91.7]**** | 34.6 | 6.3 [4.89-7.87]**** | 41.6 | 1.7 [1.50-1.99]**** | 42.9 | 1.1 [0.77-1.46]ns |
| DM-HT-CKD | 36.9 | 45.0 [35.3-55.8]**** | 42.8 | 7.8 [7.25-8.31]**** | 48.9 | 2.0 [1.93-2.15]**** | 55.5 | 1.4 [1.22-1.62]**** |
| DM-HT-CKD-smk | 42.9 | 52.2 [19.3-91.7]**** | 52.1 | 9.5 [7.39-11.5]**** | 44.7 | 1.9 [1.48-2.27]**** | 87.5 | 2.2 [1.36-2.53]** |
| DM-HT-CVD | 12.0 | 14.6 [5.07-36.6]** | 19.6 | 3.6 [2.91-4.34]**** | 34.1 | 1.4 [1.29-1.56]**** | 47.8 | 1.2 [1.06-1.40]** |

<sup>1</sup> All RRs were calculated against the population with no comorbidities in the age-group; 661,254 cases and 70,139 COVID-19 deaths with known comorbidity status, were included, as in table 1. The number of cases and deaths with each comorbidity combination and are in Table S8. \*\*\*\* p<0.0001; \*\*\* p<0.001; \*\* p<0.01; \* p<0.05; ns, not significant; Disease abbreviations as in text, additionally: DM, Diabetes Mellitus; Ob, obesity; smk, smoker.

Similar to other populations [26-31], smoking and asthma as single comorbidities, had RRd near 1 in Mexicans (Figure 6C-E, Table 1). For smoking, the RRd was lower in females, but below 1 in each gender (Table S5); and when breaking down by age groups it was below 1 only in 20-39 yo (Table 1). This is compatible with smoking’s health hazards being more notorious after chronic consumption and often observed at older ages. In the Mexican population, the estimated prevalence of current smoking is at least double than found in COVID-19 cases (Figure 5B), which could be due to underreporting in the COVID-19 dataset or be related to the low risk found. Smokers in COVID-19 cases were similar to the estimated prevalence of daily smokers in the Mexican population (Figure 5B). Details about smoking habits, were unavailable for analysis in the COVID-19 datasets. The RRd of smoking increased substantially when combined with other pre-existing diseases, even in young adults (Tables 1-2).

In summary, in Mexico the RR of pre-existing chronic diseases for COVID-19 death, depends on patient’s age and on the type and number of comorbidities. Our analysis suggests that the risk contributed by most comorbidities is large in children, young and middle-aged adults and decreases substantially with age, tending towards 1 in adults >60-yo. The frequent combinations of diabetes and other disease(s), that suggest advanced metabolic disease, increases risk of COVID-19 death more than 7-fold in adults 20-39 yo and more than 2.5-fold in 40-59 yo.

### Multivariate Correspondence Analysis and Hierarchical Clustering on Principal Components

Multivariate correspondence analysis, a statistical technique to reduce dimensions in data while preserving variation; with subsequent clustering of principal components, detected three clusters that separated COVID-19 outcomes in Mexicans (Figure S3). Favorable outcomes like no-hospitalization, no-pneumonia, and being alive at cutoff date, correlated with ages 0-39, and with having no comorbidity or presenting only asthma or smoking. At the opposite end, ages >60-yo, correlated with adverse outcomes like hospitalization, pneumonia, intubation and death, along with having ≥3 comorbidities and/or pre-existing conditions like CKD, diabetes, hypertension, CVD or COPD. A middle cluster included ages 40-59 and comorbidities like obesity and immunosuppression. These associations parallel those described in the previous section, and support the age-group partitions used for RR analyses.

### Years of life lost to premature COVID-19 mortality (YLL) during the first eight months of the outbreak in Mexico

YLL to COVID-19 were calculated using age 70 as the threshold for premature mortality, to observe distribution across genders, age-groups, and specific comorbidities. At end of study (Nov/2/2020,), of the 89,000 COVID-19 deaths with a positive SARS-CoV-2 RT-PCR in Mexico, 66.5% corresponded to individuals <70-yo and amounted to almost 880,000 YLL, with an important contribution from middle-aged males (Figure 7A, Table S8). Most YLL (69%) derived from individuals with at least one comorbidity (31.4% and 37.6% from individuals with one and more than one comorbidities, respectively); while 31% of YLL came from individuals without known comorbidity. Individuals that died from COVID-19 without know comorbity lost on average more years or life that those with comorbitiy (16.4 vs 14.2 years/individual that died under the age of 70).

**Figure 7.**
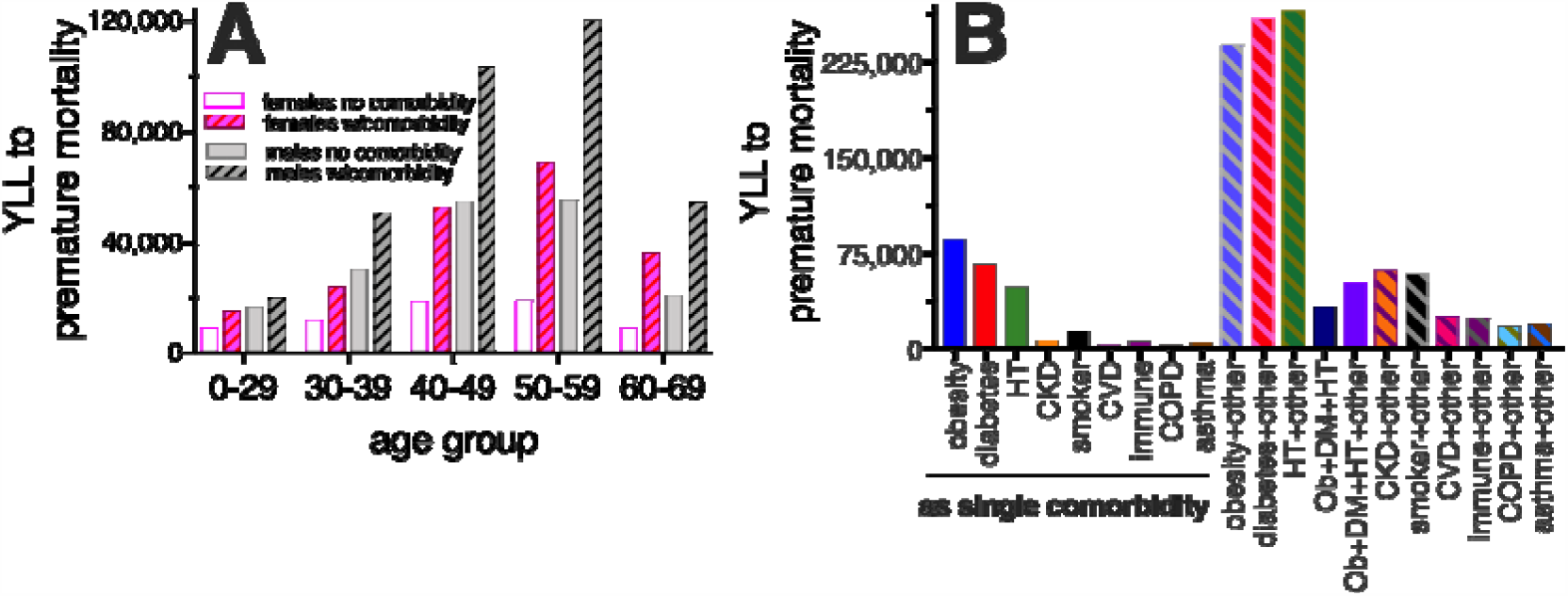
Years of life lost (YLL) due to premature COVID-19 mortality, **(A)** by age, gender and presence/absence of comorbidity; and **(B)** by specific type of comorbidity. YLL were calculated for each deceased individual <70 yo. Details in methods and Table S8. Disease abbreviations as in text; w/comorbidity, with comorbidity.

The comparison between YLL in population with and without comorbidity, suggests that in Mexico ∼38% of YLL, have been lost due to the co-occurence of COVID-19 and chronic disease (Table S8). Related to their high prevalence in the general population, obesity, diabetes and hypertension participate in the most YLL from COVID-19 in Mexico, in particular when in combination with another chronic disease (multimorbidity). These common comorbidities surpass the YLL from conditions with higher RRd, like CKD (Figure 7B). YLL calculated here are largely underestimated because not all COVID-19 deaths have been detected in Mexico, as suggested by excess mortality analyses [25], plus we excluded the >3,000 deaths determined as COVID-19 in the absence of RT-PCR. Eight months into the Mexican outbreak likely over a million years of life have been lost to COVID-19.

## Discussion

Despite its young population, Mexico is among the ten top countries in COVID-19 mortality, with ∼7.5% of world deaths. We described four components of this high mortality that deserve attention. *First*, Mexico has high COVID-19 case fatality, not in decline as in other countries, driven by low testing and high hospital-fatality. Quality and timing of medical attention await more analysis to tackle this trend [19]. *Second*, the risk contributed by comorbidities decreases with age in Mexicans, because adults >60-yo have high fatality rates even in the absence of pre-existing disease. *Third*, more than half of COVID-19 fatalities have occurred in adults younger than 65-yo, in contrast to high-income nations. This is related to a rise in COVID-19 mortality and case fatality starting at ages 45-50, due in part to chronic comorbidities in middle-aged adults in Mexico. In particular individuals that accumulate more than one chronic metabolic disease lead to a large toll of years of life lost to premature mortality from COVID-19. *Fourth*, ¼ of deaths and ∼1/3 of YLL have occurred in individuals without known comorbidity, specially males 40-60 yo.

We propose that all these observations fall on the spectrum of gradual health decline that accompanies aging. Given that a large proportion of Mexican adults are obese (36%) or overweight (39%) [13-14], environmental and genetic factors that favor metabolic disease could be contributing to early-aging in Mexicans, first driving metabolic-inflammation then leading to-, or merging with-, age-related inflammation. Both ultimately contribute to immune senescence and undermine the chances of clearing infection. Undiagnosed/subclinical metabolic disease, or being overweight, glucose-intolerant, malnourished, sarcopenic or exposed to pollutants, may contribute to adverse COVID-19 outcomes along this spectrum, by favoring lung, metabolic and immune-senescence [4-5,32-35]. Our analysis agrees with aging as a main independent risk factor for COVID-19 fatality, and with chronic diseases as important markers of aging. A shift towards middle-age distribution of COVID-19 fatalities is likely not unique to Mexico and awaits systematic exploration in other low- and middle-income populations [7].

Our analysis provides information to distinguish vulnerable subpopulations for special attention during outbreaks and for SARS-CoV-2 immunization, when available; and is compatible with the emerging knowledge about risks of comorbidity for COVID-19 [6,26-48]. Additionally, we provide new information on the age-stratified relative risk of 23 specific comorbidity-combinations that can be extrapolated to other populations. CKD has emerged as the comorbidity with the highest risk for COVID-19 death [6,26,38-41]. Common comorbidity combinations, like diabetes/hypertension with or without obesity, have RRd as high as that of CKD, related to their pathogenesis with chronic inflammation that alters the immune response to SARS-CoV-2 [42-43]. Hypertension, diabetes, COPD and CVD are markers of the aged population and contribute a low organ reserve to resist severe infection [6,26,32-35,42]. We estimate that in Mexico, up to ∼38% of YLL, have been lost due to the synergy between COVID-19 and chronic disease (Table S8), specially obesity, diabetes and hypertension (Figure 6B). Plus, our analysis suggests that advanced metabolic disease, as marked by diabetes or by multimorbidity, substantially increases the RR of COVID-19 death in adults <60-yo and could account for some of the differences in mortality between countries. In Mexico and other world regions, obesity and metabolic disease have been shifting towards disadvantaged socioeconomic groups [49] that are also more exposed to SARS-CoV-2 [50].

Importantly, our analysis highlights that during large COVID-19 outbreaks, fatalities in middle-age adults rise, making them a group of concern after elders. Mexico and USA have similar age-stratified prevalence of obesity and diabetes (Table S9) [13-14,51-53], yet Mexico has much larger COVID-19 mortality in 45-74 yo (Figure 3D). These differences can be related to the bigger toll of chronic diseases in poorer populations [52,54], and to less access to quality medical attention in Mexico, both for the control of chronic diseases and for COVID-19. In Mexico more than half of COVID-19 deaths have happened while hospitalized in non-critical areas, suggesting that the healthcare system remains stressed. Thus, our analysis is important evidence that mortality in middle aged adults can increase during large outbreaks when hospitals become saturated.

Strengths of our study include a large sample that allowed multiple age and comorbidity-combination analyses. Limitations, include lack of detail about the severity and treatment of the studied comorbidities and uncertainty about underreporting of underlying diseases. Diabetes and hypertension were found in COVID-19 cases at a similar age-stratified prevalence to the general-population’s, which doesn’t suggest underreporting; but underreporting of obesity cannot be ruled out. Overweight adults were undistinguishable in the COVID-19 dataset, which lacks body-mass-index details. Being overweight may contribute to adverse COVID-19 outcomes in a subset of the adults classified as “without comorbidity”, and should be studied.

## Conclusions

The high COVID-19 mortality in middle-aged adults in Mexico is partially explained by the synergy with chronic metabolic diseases, in particular combinations between diabetes, hypertension and obesity. Differences in age of COVID-19 mortality between the rich and the poor around the world, likely reflect health and nutrition disparities that accelerate metabolic and inflammatory aging, and economic insecurity that difficult sheltering from the virus and access to quality medical attention. Middle-aged adults, can be vulnerable during large COVID-19 outbreaks, second to the elderly, in particular where large economic and health disparities exist.

## Data Availability

All the data used in this study is hosted by the mexican government here: https://www.gob.mx/salud/documentos/datos-abiertos-152127
It is updated daily.

## Acknowledgments

We dedicate this manuscript to the Mexican health workers who have lost their lives to COVID-19 during 2020, and to the persisting efforts of those alive, who throughout the country, have compiled the data analyzed here. Through their national effort, the Mexican COVID-19 datasets currently constitute among the largest COVID-19 cohorts for open analysis in the world. LDR has support from Universidad de las Americas Puebla (UDLAP) for a Modeling and Data Visualization Laboratory.

## Ethics Approval

The analyses presented here were done with public data where patient identity was omitted, thus no ethics approval was necessary. Information was managed according to the ethical principles of the Helsinki declaration

## Funding

No external funding

## Author Contributions

conceptualization, P.C.H.; G.S.L.; L.D.R:; methodology, P.C.H. and L.D.R.; formal analysis, P.C.H.; F.R.P, L.D.R. and F.S.J.; investigation, P.C.H.; data curation, L.D.R and F.R.P.; writing—original draft preparation, P.C.H. and G.S.L.; writing—review and editing, P.C.H. and L.D.R.; supervision, P.C.H. All authors have read and agreed to the published version of the manuscript.

## Competing interests and Conflicts of Interest

None to declare.

## Supplementary Materials

**Figure S1** Combinations of 2 comorbidities in COVID-19 deaths.

**Figure S2** Combinations of 3 or more comorbidities in COVID-19 deaths.

**Figure S3** Correspondence Analyses and Hierarchical Clustering of Principal Components (HCPC) of the Mexican COVID-19 datasets

**Table S1**. Characteristics of confirmed COVID-19 cases and deaths in Mexico until Aug/06/2020 (first cutoff).

**Table S2**. Characteristics of confirmed COVID-19 cases and deaths in Mexico until Sept/12/2020 (second cutoff).

**Table S3**. Characteristics of confirmed COVID-19 cases and deaths in Mexico until Nov/2/2020 (end of study).

**Table S4**. Age of COVID-19 cases and deaths in Mexico subdivided by the presence/absence of comorbidity, until Aug/06/2020 (first cutoff).

**Table S5**. Frequency, CFR and RRd of specific comorbidities by age group, in COVID-19 in Mexico until August/6/2020 (first cutoff).

**Table S6**. Frequency, CFR and RRd of specific comorbidities by age group, in COVID-19 in Mexico until August/6/2020 (first cutoff).

**Table S7**. Frequency of the 23 most common comorbidity combinations by age group, in Mexico until Sept/12/2020 (second cutoff).

**Table S8**. Years of life lost to premature COVID-19 mortality (YLL) in Mexico until end of study (Nov/2/2020), subdivided by gender and presence/absence of comorbidity.

**Table S9**. Age stratified comparison of diabetes and obesity prevalence in Mexico *vs* the USA, as reported by health authorities.

## Notes

### Competing Interest Statement

The authors have declared no competing interest.

### Funding Statement

No external funding was received.

### Author Declarations

The authors are researchers at Instituto Mexicano del Seguro Social, that has ethics committees supervised by the federal authority in the country (Mexico). The manuscript analyzes datasets that are publicly available and where no individual patient information is distinguishable, thus the research is exempt from informed consent. This manuscript has been submitted for peer review and publication to the Journal of Global Health.

### Summary of Updates

Improved compression of the figures to enhance its appearance.

## References

1. Chen N, Zhou M, Dong X, Qu J, Gong F, Han Y, et al. Epidemiological and clinical characteristics of 99 cases of 2019 novel coronavirus pneumonia in Wuhan, China: a descriptive study. Lancet. 2020;395:507–13.

2. CC-19 Response Team. Severe Outcomes Among Patients with Coronavirus Disease 2019 (COVID-19) — United States, February 12–March 16, 2020. MMWR 2020;69:343–346.

3. Onder G, Rezza G, Brusaferro S. Case-Fatality Rate and Characteristics of Patients Dying in Relation to COVID-19 in Italy. JAMA. 2020;323:1775–1776.

4. Shahid Z, Kalayanamitra R, McClafferty B, Kepko D, Ramgobin D, Patel R, et al. COVID-19 and Older Adults: What We Know. J Am Geriatr Soc. 2020;68:926–9. doi:10.1111/jgs.16472.

5. Bonanad C, García BS, Tarazona SF, Sanchis J, Bertomeu GV, Fácila L, et al. The Effect of Age on Mortality in Patients With COVID-19: A Meta-Analysis With 611,583 Subjects. JAMDA. 2020;21:915–918. doi:10.1016/j.jamda.2020.05.045

6. Ssentongo P, Ssentongo AE, Heilbrunn ES, Ba DM, Chinchilli VM. Association of cardiovascular disease and 10 other pre-existing comorbidities with COVID-19 mortality: A systematic review and meta-analysis. PLOS ONE. 2020;15:e0238215. doi:10.1371/journal.pone.0238215

7. Ioannidis JPA, Axfors C, Contopoulos-Ioannidis DG. Population-level COVID-19 mortality risk for non-elderly individuals overall and for non-elderly individuals without underlying diseases in pandemic epicenters. Environ Res. 2020;188:109890. doi:10.1016/j.envres.2020.109890

8. Population Pyramids of the World from 1950 to 2100. Database: Population pyramid. Available at: https://www.populationpyramid.net/mexico/2020/. Accessed 12 October 2020.

9. International Institute for Applied Systems Analysis (IIASA). 2018. Aging Demographic Data Sheet 2018. Available at: https://iiasa.ac.at/web/home/research/researchPrograms/WorldPopulation/PublicationsMediaCoverage/ModelsData/AgingDemDataSheet2018_web.pdf. Accessed 12 October 2020.

10. Our World in Data. Coronavirus Source Data. Available at: https://ourworldindata.org/coronavirus-source-data. Accessed 12 October 2020.

11. Suárez V, Suarez QM, Oros RS, Ronquillo DE. Epidemiology of COVID-19 in Mexico: From the 27th of February to the 30th of April 2020. Rev Clin Esp. 2020;220:463–471.

12. INEGI. Características de las defunciones registradas en México durante 2019. Comunicado de prensa núm. 480/20. 29 de octubre de 2020. Available online: https://www.inegi.org.mx/contenidos/saladeprensa/boletines/2020/EstSociodemo/DefuncionesRegistradas2019.pdf (accessed on 10 November 2020)

13. Encuesta Nacional de Salud y Nutrición. Encuesta Nacional de Salud y Nutrición presentación de resultados. Available at: https://ensanut.insp.mx/encuestas/ensanut2018/doctos/informes/ensanut_2018_presentacion_resultados.pdf. Accessed 12 October 2020.

14. Shamah-Levy T, Vielma-Orozco E, Heredia-Hernández O, Romero-Martínez M, Mojica-Cuevas J, Cuevas-Nasu L, et al. Encuesta Nacional de Salud y Nutrición 2018-19: Resultados Nacionales. Cuernavaca, México: Instituto Nacional de Salud Pública, 2020.

15. Gobierno de México. Datos Abiertos Bases Históricas: Secretaría de Salud Available at: http://www.gob.mx/salud/documentos/datos-abiertos-bases-historicas-direccion-general-de-epidemiologia. Accessed 12 October 2020.

16. Denova GE, Lopez-Gatell H, Alomia ZJL, López RR, Zaragoza JCA, Leal DD, et al. The Association of Obesity, Type 2 Diabetes, and Hypertension with Severe Coronavirus Disease 2019 on Admission Among Mexican Patients. Obesity (Silver Spring). 2020;28:1826–1832. doi:10.1002/oby.22946

17. Lê S, Josse J, Husson F, FactoMineR: An R Package for Multivariate Analysis. J Stat Softw. 2008; 25: 1–18. doi:10.18637/jss.v025.i01

18. Kassambara A. Practical Guide To Principal Component Methods in R (Multivariate Analysis); 2 ISBN-13: 978-1975721138; 2017

19. Ñamendys-Silva SA, Gutiérrez VA, Romero GJP. Hospital mortality in mechanically ventilated COVID-19 patients in Mexico. Intensive Care 2020; 46(11); 2086–2088. doi:doi.org/10.1007/s00134-020-06256-3

20. Sorci G, Faivre B, Morand S. Explaining among-country variation in COVID-19 case fatality rate. Sci Rep 2020;10:18909. Doi: 10.1038/s41598-020-75848-2

21. EPICENTRO L’epidemiologia per la sanità publica Istituto Superiore di Sanità. Caratteristiche dei pazienti deceduti positivi all’infezione da SARS-CoV-2 in Italia. Avalible online: https://www.epicentro.iss.it/coronavirus/sars-cov-2-decessi-italia (accessed on 3 November 2020).

22. Center of Disease Control and Prevention. Provisional COVID-19 Death Counts by Sex, Age, and State. Avalible online: https://data.cdc.gov/NCHS/Provisional-COVID-19-Death-Counts-by-Sex-Age-and-S/9bhg-hcku (Accessed on 31 October 2020).

23. Office for National Statics. Deaths registered weekly in England and Wales, provisional: week ending 23 October 2020. Avalible online:https://www.ons.gov.uk/peoplepopulationandcommunity/birthsdeathsandmarriages/deaths/bulletins/deathsregisteredweeklyinenglandandwalesprovisional/weekending23october2020 (accessed on 31 October 2020)

24. Laxminarayan R, Wahl B, Dudala SR, Gopal K, Mohal BC, Neelima S. Epidemiology and transmission dynamics of COVID-19 in two Indian states. Science. 2020;370:691–697.

25. Grupo interinstitucional para la estimación del exceso de mortalidad por todas las causas. Boletín estadístico sobre exceso de mortalidad por todas las causas durante la emergencia por COVID-19. México, 2020;1 [Internet] [cited 2020 Nov 12] Spanish Available from: https://www.insp.mx/micrositio-covid-19/boletin-estadistico-sobre-exceso-de-mortalidad-por-todas-las-causas-durante-la-emergencia-por-covid-19-numero-2.

26. Williamson EJ. Walker AJ, Bhaskaran K, Bacon S, Bates C, Morton CE, et al. Factors associated with COVID-19-related death using OpenSAFELY. Nature. 2020;584:430–436.

27. González RJ, Navarro LC, López NE, López NA, Jiménez DL, Navarro LJD, et al. Systematic Review and Meta-Analysis of Hospitalised Current Smokers and COVID-19. Int. J. Environ. Res. Public Health 2020;17:7394. doi:10.3390/ijerph17207394

28. Lippi G, Henry BM. Active smoking is not associated with severity of coronavirus disease 2019 (COVID-19). Eur J Intern Med. 2020;75:107–108. doi:10.1016/j.ejim.2020.03.014.

29. Patanavanich R, Glantz SA. Smoking Is Associated With COVID-19 Progression: A Meta-analysis. Nicotine Tob Res. 2020;22:1653–1656. doi:10.1093/ntr/ntaa082.

30. Broadhurst R, Peterson R, Wisnivesky JP, Federman A, Shanta M, Zimmer, et al. Asthma in COVID-19 Hospitalizations: An Overestimated Risk Factor? Annals of the American Thoracic Society. 2020. doi:10.1513/AnnalsATS.202006-613RL

31. Matsumoto K, Saito H. Does asthma affect morbidity or severity of COVID-19? J Allergy Clin Immunol. 2020;146:55–57. doi:10.1016/j.jaci.2020.05.017.

32. Mueller AL, McNamara MS, Sinclair DA. Why does COVID-19 disproportionately affect older people? Aging. 2020;12:9959–9981. doi:10.18632/aging.103344

33. Mallapaty S. The coronavirus is most deadly if you are older and male — new data reveal the risks. Nature. 2020;585:16–17. doi:10.1038/d41586-020-02483-2.

34. Ma Y, Hou L, Yang X, Huang Z, Yang X, Zhao N, et al. The association between frailty and severe disease among COVID-19 patients aged over 60 years in China: a prospective cohort study. BMC Med, 2020;18:274. doi:10.1186/s12916-020-01761-0

35. Bonafè M, Prattichizzo F, Giuliani A, Storci G, Sabbatinelli J, Olivieri F. Inflamm-aging: Why older men are the most susceptible to SARS-CoV-2 complicated outcomes. Cytokine Growth Factor Rev. 2020; 53: 33–37. doi:10.1016/j.cytogfr.2020.04.005

36. Carrillo VMF, Salinas EG, García PC, Gutiérrez RLM, Parra RL. Early estimation of the risk factors for hospitalization and mortality by COVID-19 in Mexico. PLOS One. 2020; 15(9): e0238905. doi:10.1371/journal.pone.0238905

37. Hernández GDR, González BMA, Romo DDK, Lima MR, Hernández VIA, Lumbreras GM, et al. Increased Risk of Hospitalization and Death in Patients with COVID-19 and Pre-existing Noncommunicable Diseases and Modifiable Risk Factors in Mexico. Arch Med Res. 2020; 51 (7); 683–689. doi:10.1016/j.arcmed.2020.07.003

38. Gansevoort RT, Hilbrands LB. CKD is a key risk factor for COVID-19 mortality. Nat Rev Nephrol 2020; 395: 497–506. doi:10.1007/s10900-020-00920-x

39. Henry BM, Lippi G. Chronic kidney disease is associated with severe coronavirus disease 2019 (COVID-19) infection. Int Urol Nephrol. 2020; 52:1193–1194. doi:10.1007/s11255-020-02451-9.

40. Ajaimy M, Melamed ML. COVID-19 in Patients with Kidney Disease. CJASN 2020; 15: 1087–1089. doi:10.2215/CJN.09730620

41. Cheng Y, Luo R, Wang K, Zhang M, Wang Z, Dong L, et al. Kidney disease is associated with in-hospital death of patients with COVID-19. Kidney Int. 2020; 97: 829–838. doi:10.1016/j.kint.2020.03.005

42. De Almeida PB, Dualib PM, Zajdenverg L, Rodrigues DJ, Días SF, Rodacki M, et al. Severity and mortality of COVID 19 in patients with diabetes, hypertension and cardiovascular disease: a meta-analysis. Diabetol Metab Syndr 2020; 12:75. doi:10.1186/s13098-020-00586-4

43. Apicella M, Campopiano MC, Mantuano M, Mazoni L, Coppelli A, Del Prato S. COVID-19 in people with diabetes: understanding the reasons for worse outcomes. Lancet Diabetes Endocrinol. 2020; 8:782–792. doi:10.1016/S2213-8587(20)30238-2

44. Pasquarelli NG, Braz MHA, Faria SS, Santos IO, Kobinger GP, Magalhães KG. Hypercoagulopathy and adipose tissue exacerbated inflammation may explain higher mortality in COVID-19 patients with obesity. Front Endocrinol (Lausanne). 2020; 11: 530. doi:10.3389/fendo.2020.00530

45. Tartof S, Qian L, Hong V, Wei R, Nadjafi RF, Fischer H, et al. Obesity and mortality among patients diagnosed with COVID-19: Results from an integrated health care organization. Ann Int Med 2020; M20–3742. doi:10.7326/M20-3742

46. Nakeshbandi M, Maini R, Daniel P, Rosengarten S, Parmar P, Wilson C, et al. The impact of obesity on COVID-19 complications: a retrospective cohort study. Int J Obes 2020; 44, 1832–1837. doi:10.1038/s41366-020-0648-x

47. Popkin BM, D. S, Green WD, Beck MA, Algaith T, Herbst CH, et al. Individuals with obesity and COVID-19: A global perspective on the epidemiology and biological relationships. Obes Rev. 2020; 21:e13128. doi:10.1111/obr.13128

48. Parra BGM, López VN, Parra BFE. Clinical characteristics and risk factors for mortality of patients with COVID-19 in a large data set from Mexico. Ann.Epidemiol. 2020; doi:10.1016/j.annepidem.2020.08.005

49. Jiwani SS, Carrillo-Larco RM, Hernández-Vásquez A, Barrientos-Gutiérrez T, Basto-Abreu A, Gutierrez L et al. The shift of obesity burden by socioeconomic status between 1998 and 2017 in Latin America and the Caribbean: a cross-sectional series study Lancet Global Health. 2019;7:e1644-e1654. 10.1016/S2214-109X(19)30421-8

50. Chang S, Pierson E, Kho PW, Gerardin J, Redbird B, Grusky D et al. Mobility network models of COVID-19 explain inequities and inform reopening. Nature. 2020.10.1038/s41586-020-2923-3

51. Hales MC, Carroll DM, Fryar DC, Ogden LC. Prevalence of Obesity Among Adults and Youth: United States, 2015–2016. NCHS Data Brief. 2017; 288.

52. Fryar DC, Carroll DM, Ogden LC. Prevalence of Overweight, Obesity, and Severe Obesity Among Children and Adolescents Aged 2–19 Years: United States, 1963–1965 Through 2015–2016. Avalible at: https://www.cdc.gov/nchs/data/hestat/obesity_child_15_16/obesity_child_15_16.html Accessed 11 Novembrer 2020.

53. National Diabetes Statistics Report 2020. Estimates of Diabetes and Its Burden in the United States Avalible at: https://www.cdc.gov/diabetes/pdfs/data/statistics/national-diabetes-statistics-report.pdf Accessed 11 November 2020

54. Dai H, Alsalhe TA, Chalghaf N, Riccò M, Bragazzi NL, Wu J. The global burden of disease attributable to high body mass index in 195 countries and territories, 1990-2017: An analysis of the Global Burden of Disease Study. PLoS Med. 2020;17:e1003198.

