## Supplemental Material for "The role of metabolic comorbidity in COVID-19 mortality of middle-aged adults. The case of Mexico"

**Table S1**. Characteristics of confirmed COVID-19 cases and deaths in Mexico until Aug/06/2020 (first cutoff).

| | Cases | | Deaths | | % | | Mortality |
|---|---|---|---|---|---|---|---|
| | n (%) | Age | n (%) | Age | mCFR | $RR_d$ [95% CI] [a] | (100,000 inh.) [b] |
| Males | 245,583 (53.1) | 45.8±16.5 | 32,707 (64.7) | 61.1±14.0 | 13.32 | 1.62 [1.60-1.65]**** | 51.8 |
| Females | 217,107 (46.9) | 44.4±16.4 | 17,810 (35.3) | 62.8±14.3 | 8.20 | 0.62 [0.61-0.63]**** | 27.0 |
| Total | 462,690 (100) | 45.1±16.4 | 50,517 (100) | 61.7±14.2 | 10.92 | - | 39.1 |
| *Age group (years old)* | | | | | | | |
| 0-1 | 1,596 (0.3) | - | 78 (0.2) | - | 4.89 | 0.45 [0.36-0.55]**** | |
| 2-9 | 3,216 (0.7) | - | 44 (0.1) | - | 1.37 | 0.13 [0.09-0.17]**** | 0.5 |
| 10-19 | 10,992 (2.4) | - | 85 (0.2) | - | 0.77 | 0.07 [0.06-0.09]**** | 0.3 |
| 20-29 | 66,745 (14.4) | - | 614 (1.2) | - | 0.92 | 0.07 [0.07-0.08]**** | 2.8 |
| 30-39 | 103,293 (22.3) | - | 2,344 (4.6) | - | 2.27 | 0.17 [0.16-0.18]**** | 12.4 |
| 40-49 | 101,647 (22.0) | - | 6,377 (12.6) | - | 6.27 | 0.51 [0.50-0.53]**** | 38.6 |
| 50-59 | 83,567 (18.1) | - | 11,688 (23.1) | - | 13.99 | 1.37 [1.34-1.39]**** | 93.2 |
| 60-69 | 52,327 (11.3) | - | 13,827 (27.4) | - | 26.42 | 2.96 [2.91-3.01]**** | 171.1 |
| 70-79 | 27,270 (5.9) | - | 10,254 (20.3) | - | 37.60 | 4.07 [3.99-4.14]**** | 252.1 |
| 80-89 | 10,379 (2.2) | - | 4,521 (8.9) | - | 43.56 | 4.28 [4.18-4.39]**** | 274.6 |
| ≥90 | 1,658 (0.4) | - | 685 (1.4) | - | 41.31 | 3.82 [3.60-4.05]**** | 181.7 |
| Total | 462,690 (100) | | 50,517 (100) | | 10.92 | - | |
| *Clinical condition* | | | | | | | |
| Not hospitalized | 338,693 (73.2) | 41.2±14.7 | 5696 (11.3) | 60.8±14.4 | 1.68 | 0.05 [0.045-0.048]**** | ND |
| Hospitalized | 123,997 (26.8) | 56.0±16.1 | 44,821 (88.7) | 61.8±14.1 | 36.15 | 21.5 [20.93-22.08]**** | ND |
| Unknown condition | 116 (0.03) | 55.2±16.5 | 69 (0.1) | 59.4±17.1 | 59.48 | - | ND |
| Not critical[c] | 107,145 (23.2) | 55.8±16.1 | 34,749 (68.8) | 62.4±14.0 | 32.43 | 19.3 [18.77-19.82]**** | ND |
| Critical | 16,736 (3.6) | 56.9±16.4 | 10,003 (19.8) | 59.9±14.4 | 59.77 | 35.5 [34.54-36.57]**** | ND |
| AMV | 11,928 (2.6) | 58.0±15.3 | 8,642 (17.1) | 59.7±14.2 | 72.45 | 43.1 [41.89-44.30]**** | ND |
| Confirmed Pneumonia | 94,133 (20.3) | 55.9±15.8 | 37,699 (74.6) | 61.7±14.1 | 40.05 | 11.5 [11.30-11.73]**** | ND |
| *Number of comorbidities* | | | | | | | |
| 0 | 249,308 (53.9) | 40.2±15.1 | 13,467 (26.7) | 59.6±15.0 | 5.40 | 0.31 [0.30-0.32]**** | ND |
| 1 | 122,233 (26.4) | 47.4±15.8 | 15,056 (29.9) | 61.3±14.6 | 12.32 | 2.28 [2.23-2.33]**** | ND |
| 2 | 59,246 (12.8) | 54.4±15.3 | 12,513 (24.8) | 63.2±13.3 | 21.12 | 3.91 [3.82-4.00]**** | ND |
| ≥3 | 30,016 (6.5) | 58.6±13.9 | 9,146 (18.1) | 63.6±12.6 | 30.47 | 5.64 [5.51-5.78]**** | ND |
| unknown | 1,887 (0.4) | 47.4±16.6 | 335 (0.7) | 59.0±14.8 | 17.75 | - | ND |
| Total | 462,690 (100) | | 50,517 (100) | | 10.92 | - | |
| Total with known comorbidity status | 460,803 (99.6) | | 50,182 (99.3) | | 10.89 | - | |

[a]RR of death was calculated for males vs females; pregnant females vs 20-39 yo non-pregnant females; indigenous language speakers vs non-speakers; each age group vs the rest of the population; not hospitalized vs hospitalized and *vice versa*; other clinical conditions vs not hospitalized; having vs not having confirmed pneumonia; having no comorbidity (0) vs having some comorbidity (≥1); having 1, 2, ≥3 comorbidities vs no comorbidity. **** $p < 0.0001$, *** $p < 0.001$, CI= Confidence interval.

[b] Mortality corresponds to COVID-19 deaths until Aug 06, 2020 and has increased since. ND=not determined.

[c] "Not critical" means that the patient was not treated in the ICU (Intensive Care Unit) and was not intubated outside the ICU up until the cutoff date (Aug/06/2020). "Critical" patients were those treated in the ICU or that required intubation and mechanical ventilation even if they were not in the ICU.

**Table S2**. Characteristics of confirmed COVID-19 cases and deaths in Mexico until Sept/12/2020 (second cutoff).

| | Cases | | Deaths | | % | | Mortality |
|---|---|---|---|---|---|---|---|
| | n (%) | Age | n (%) | Age | mCFR | RR [95% CI] [a] | (100,000 inh.) [b] |
| Males | 346,464 (52.2) | 45.5± 16.7 | 45,413 (64.3) | 61.7± 14.1 | 13.11 | 1.65 [1.63-1.68]$^{****}$ | 72.0 |
| Females | 317,509 (47.8) | 44.1± 16.5 | 25,191 (35.7) | 63.3± 14.2 | 7.93 | 0.61 [0.60-0.61]$^{****}$ | 38.2 |
| Total | 663,973 (100) | 44.8± 16.6 | 70,604 (100) | 62.2± 14.2 | 10.63 | - | 54.8 |
| *Age group (years old)* | | | | | | | |
| 0-1 | 2,233 (0.3) | - | 107 (0.2) | - | 4.79 | 0.45 [ 0.37-0.54]$^{****}$ | |
| 2-9 | 5,032 (0.8) | - | 52 (0.1) | - | 1.03 | 0.10 [ 0.07-0.13]$^{****}$ | |
| 10-19 | 17,926 (2.7) | - | 12 (0.2) | - | 0.67 | 0.06 [ 0.05-0.07]$^{****}$ | 0.5 |
| 20-29 | 100,792 (15.2) | - | 851 (1.2) | - | 0.84 | 0.07 [ 0.06-0.07]$^{****}$ | 3.9 |
| 30-39 | 147,859 (22.3) | - | 3,081 (4.4) | - | 2.08 | 0.16 [ 0.15-0.17]$^{****}$ | 16.4 |
| 40-49 | 143,051 (21.5) | - | 8,468 (12.0) | - | 5.92 | 0.50 [ 0.49-0.51]$^{****}$ | 51.3 |
| 50-59 | 117,449 (17.7) | - | 15,836 (22.4) | - | 13.48 | 1.35 [ 1.32-1.37]$^{****}$ | 126.4 |
| 60-69 | 73,679 (11.1) | - | 19,529 (27.7) | - | 26.51 | 3.06 [ 3.02-3.11]$^{****}$ | 243.2 |
| 70-79 | 38,765 (5.8) | - | 14,915 (21.1) | - | 38.48 | 4.32 [ 4.26-4.38]$^{****}$ | 366.8 |
| 80-89 | 14,816 (2.2) | - | 6,660 (9.4) | - | 44.95 | 4.56 [ 4.48-4.65]$^{****}$ | 404.6 |
| ≥90 | 2,371 (0.4) | - | 984 (1.4) | - | 41.50 | 3.94 [3.76-4.14]$^{****}$ | 261.1 |
| Total | 663,973 (100) | | 70,604 (100) | | 10.63 | - | |
| *Clinical condition* | | | | | | | |
| Not hospitalized | 500,193 (75.3) | 41.0± 14.9 | 8,110 (11.5) | 61.4± 14.4 | 1.62 | 0.04 [0.04-0.04]$^{****}$ | ND |
| Hospitalized | 163,780 (24.7) | 56.5± 16.3 | 62,494 (88.5) | 62.4± 14.1 | 38.16 | 23.5 [ 23.01-24.07]$^{****}$ | ND |
| Unknown condition | 152 (0.02) | 56.2±15.3 | 74 (0.1) | 59.5± 16.1 | 48.68 | - | ND |
| Not critical[c] | 127,337 (19.2) | 55.9± 16.4 | 36,644 (51.9) | 63.4± 14.1 | 28.78 | 17.8 [17.34-18.17]$^{****}$ | ND |
| Critical | 36,291 (5.5) | 58.4± 15.7 | 25,776 (36.5) | 60.9± 14.0 | 71.03 | 43.8 [42.83-44.81]$^{****}$ | ND |
| AMV | 29,850 (4.5) | 59.3± 14.9 | 24,065 (34.1) | 60.8± 13.9 | 80.62 | 49.7 [48.63-50.84]$^{****}$ | ND |
| Confirmed Pneumonia | 125,765 (18.9) | 56.4± 16.0 | 52,398 (74.2) | 62.2± 14.1 | 41.66 | 12.3 [ 12.12-12.51]$^{****}$ | ND |
| *Number of comorbidities* | | | | | | | |
| 0 | 362,425 (54.6) | 39.8± 15.3 | 18,553 (26.3) | 60.1± 15.0 | 5.12 | 0.30 [0.29-0.30]$^{****}$ | ND |
| 1 | 174,141 (26.2) | 47.3± 16.0 | 20,975 (29.7) | 61.9± 14.6 | 12.04 | 2.35 [2.31-2.40]$^{****}$ | ND |
| 2 | 83,165 (12.5) | 54.5± 15.4 | 17,703 (25.1) | 63.7± 13.3 | 21.29 | 4.16 [4.08-4.24]$^{****}$ | ND |
| ≥3 | 41,523 (6.3) | 58.8± 14.0 | 12,908 (18.3) | 64.0± 12.7 | 31.09 | 6.07 [5.95-6.20]$^{****}$ | ND |
| unknown | 2,719 (0.4) | 46.9± 17.0 | 465 (0.7) | 59.2± 15.5 | 17.10 | - | ND |
| Total | 663,973 (100) | | 70,604 (100) | | 10.63 | - | |
| Total with known comorbidity status | 661,254 (99.6) | | 70,139 (99.3) | | 10.61 | - | |

[a]RRd were calculated as for Supplementary Table 1. *** $p < 0.0001$, ** $p < 0.001$, CI= Confidence interval.

[b] Mortality depicted here corresponds to COVID-19 deaths until Sep 12, 2020 and has increased since.

[c] "Not critical" means that the patient was not treated in the ICU (Intensive Care Unit) and was not intubated outside the ICU up until the cutoff date (Sep/12/2020). "Critical" patients were those treated in the ICU or that required intubation and mechanical ventilation even if they were not in the ICU.

**Table S3.** Characteristics of confirmed COVID-19 cases and deaths in Mexico until Nov/2/2020 (end of study).

| | Cases | | Deaths | | % | | Mortality |
|---|---|---|---|---|---|---|---|
| | n (%) | Age | n (%) | Age | mCFR | $RR_d$ [95% CI] [a] | (100,000 inh.) [b] |
| Males | 465,079 (51.4) | 45.0±16.9 | 56,916 (63.8) | 62.1±14.2 | 12.24 | 1.67 [1.65-1.69]**** | 90.2 |
| Females | 440,500 (48.6) | 43.7±16.5 | 32,251 (36.2) | 63.5±14.2 | 7.32 | 0.60 [0.59-0.61]**** | 49.0 |
| Total | 905579 (100) | 44.4±16.7 | 89,167 (100) | 62.6±14.2 | 9.85 | - | 69.2 |
| *Age group (years old)* | | | | | | | |
| 0-1 | 2,943 (0.3) | - | 132 (0.1) | - | 4.49 | 0.45 [0.38-0.54]**** | - |
| 2-9 | 7,172 (0.8) | - | 67 (0.1) | - | 0.93 | 0.09 [0.07-0.12]**** | - |
| 10-19 | 27,492 (3.0) | - | 156 (0.2) | - | 0.57 | 0.06 [0.05-0.07]**** | 0.7 |
| 20-29 | 145,299 (16.0) | - | 1,063 (1.2) | - | 0.73 | 0.06 [0.06-0.07]**** | 4.9 |
| 30-39 | 201,362 (22.2) | - | 3,744 (4.2) | - | 1.86 | 0.15 [0.15-0.16]**** | 19.9 |
| 40-49 | 192,365 (21.2) | - | 10,314 (11.6) | - | 5.36 | 0.49 [0.48-0.49]**** | 62.4 |
| 50-59 | 157,082 (17.3) | - | 19,546 (21.9) | - | 12.44 | 1.34 [1.32-1.36]**** | 156.0 |
| 60-69 | 97,716 (10.8) | - | 24,709 (27.7) | - | 25.29 | 3.17 [3.13-3.21]**** | 307.7 |
| 70-79 | 51,337 (5.7) | - | 19,322 (21.7) | - | 37.64 | 4.60 [4.54-4.66]**** | 475.1 |
| 80-89 | 19,642 (2.2) | - | 8,787 (9.9) | - | 44.74 | 4.93 [4.85-5.01]**** | 533.8 |
| $\geq$90 | 3,169 (0.3) | - | 1,327 (1.5) | - | 41.87 | 4.30 [4.13-4.48]**** | 352.1 |
| Total | 905,579 (100) | | 89,167 (100) | | 9.85 | - | |
| *Clinical condition* | | | | | | | |
| Not hospitalized | 701,279 (77.4) | 40.7±15.0 | 9,690 (10.9) | 61.7±14.4 | 1.38 | 0.04 [0.03-0.04]**** | ND |
| Hospitalized | 204,300 (22.6) | 56.8±16.4 | 79,477 (89.1) | 62.7±14.2 | 38.90 | 28.2 [27.58-28.74]**** | ND |
| Unknown condition | 589 (0.07) | 59.7±15.1 | 495 (0.6) | 60.7±15.1 | 84.04 | - | ND |
| Not critical[c] | 161,850 (17.9) | 56.3±16.5 | 49,282 (55.3) | 63.7±14.1 | 30.45 | 22.0 [21.58-22.51]**** | ND |
| Critical | 41,853 (4.6) | 58.6±15.7 | 29,692 (33.3) | 61.1±14.1 | 70.94 | 51.3 [50.29-52.42]**** | ND |
| AMV | 33,738 (3.7) | 59.4±14.9 | 27,381 (30.7) | 60.9±14.0 | 81.16 | 58.7 [57.55-59.95]**** | ND |
| Confirmed Pneumonia | 155,429 (17.2) | 56.7±16.1 | 65,412 (73.4) | 62.6±14.1 | 42.08 | 13.3 [13.11-13.47]**** | ND |
| *Number of comorbidities* | | | | | | | |
| 0 | 505,783 (55.9) | 39.4±15.3 | 23,174 (26.0) | 60.6±15.1 | 4.58 | 0.28 [0.27-0.28]**** | ND |
| 1 | 232,890 (25.7) | 47.0±16.1 | 26,244 (29.4) | 62.2±14.7 | 11.27 | 2.46 [2.42-2.50]**** | ND |
| 2 | 108,992 (12.0) | 54.4±15.5 | 22,386 (25.1) | 64.0±13.4 | 20.54 | 4.48 [4.41-4.56]**** | ND |
| $\geq$3 | 53,979 (6.0) | 59.0±14.0 | 16,806 (18.8) | 64.3±12.6 | 31.13 | 6.80 [6.68-6.92]**** | ND |
| unknown | 3,935 (0.4) | 45.8±16.9 | 557 (0.6) | 60.1±15.6 | 14.16 | - | ND |
| Total | 905,579 (100) | | 89,167 (100) | | 9.85 | - | |
| Total with known comorbidity status | 901,644 (99.6) | | 88,610 (99.4) | | 9.83 | - | |

[a] $RR_d$ was calculated as in table 1. *** $p < 0.0001$, ** $p < 0.001$, CI= Confidence interval.

[b] Mortality depicted here corresponds to COVID-19 deaths until Oct 4, 2020 and has increased since.

[c] "Not critical" means that the patient was not treated in the ICU (Intensive Care Unit) and was not intubated outside the ICU up until the cutoff date (Oct/4/020). "Critical" patients were those treated in the ICU or that required intubation and mechanical ventilation even if they were not in the ICU.

**Table S4.** Age of COVID-19 cases and deaths in Mexico subdivided by the presence/absence of comorbidity, until Aug/06/2020 (first cutoff).

| | Age of Cases [a] | | | Age of deaths [a] | | |
|---|---|---|---|---|---|---|
| | without comorbidity | with comorbidity[b] | *p* [c] | without comorbidity | with comorbidity[b] | *p* [d] |
| *Clinical conditions (all genders)* | | | | | | |
| Not hospitalized | 38.0 ± 13.7 | 46.3 ± 14.8 | **** | 58.8 ± 15.4 | 61.7 ± 13.9 | **** |
| Hospitalized | 51.2 ± 17.1 | 58.6 ± 15.1 | **** | 59.7 ± 15.0 | 62.7 ± 13.8 | **** |
| Not critical [e] | 51.1 ± 16.9 | 58.5 ± 15.1 | **** | 60.5 ± 14.7 | 63.2 ± 13.8 | **** |
| Critical [e] | 52.2 ± 18.8 | 58.9 ± 14.9 | **** | 57.1 ± 15.7 | 61.0 ± 13.8 | **** |
| Intubated [e] | 54.3 ± 17.1 | 59.6 ± 14.3 | **** | 56.7 ± 15.3 | 60.9 ± 13.6 | **** |
| Confirmed Pneumonia | 51.0 ± 16.6 | 58.5 ± 14.9 | **** | 59.7 ± 14.9 | 62.5 ± 13.8 | **** |
| *Genders* | | | | | | |
| Males | 41.3 ± 15.6 | 51.0 ± 16.1 | **** | 59.3 ± 14.7 | 62.0 ± 13.8 | **** |
| Females | 39.0 ± 14.7 | 51.0 ± 16.0 | **** | 60.4 ± 15.9 | 63.5 ± 13.8 | **** |
| Total | 40.2 ± 15.2 | 51.0 ± 16.1 | **** | 59.6 ± 15.0 | 62.6 ± 13.8 | **** |

[a] Ages and standard deviation correspond to cases and deaths with known comorbidity status.
[b] These patients had the diagnosis of at least one chronic comorbidity.
[c] Two-tailed comparison of the ages of cases with *vs* without comorbidity;
[d] Two tailed comparison of the ages of individuals that died, with *vs* without comorbidity.
**** $p > 0.0001$; ns, not significant.
[e] "Not critical" refers to patients that were not in the ICU or intubated until cutoff date. "Critical" refers to patients treated in ICU or intubated. For simplicity, only the intubated patients, and not all categories of the critical patients, are shown.
*Indigenous LS*, Indigenous language speakers

**Table S5.** Frequency, CFR and RR of COVID-19 death (RRd) of specific chronic comorbidities, by gender, in Mexico, until Aug/06/2020 (first cutoff).

| | Cases n (%) | Deaths n (%) | As a single comorbidity %m CFR | As a single comorbidity $RR_d$ [95% CI][a] | In combination with one other comorbidity %m CFR | In combination with one other comorbidity $RR_d$ [95% CI][a] | In combination with ≥2 other comorbidities %m CFR | In combination with ≥2 other comorbidities $RR_d$ [95% CI][a] |
|---|---|---|---|---|---|---|---|---|
| Males | | | | | | | | |
| Obesity | 44,118 (38.6) | 7,342 (32.3) | 10.9 | 1.5 [1.39-1.52]**** | 16.6 | 2.2 [2.13-2.31]**** | 29.5 | 4.0 [3.81-4.10]**** |
| Diabetes | 39,594 (34.6) | 11,353 (49.9) | 22.1 | 3.0 [2.85-3.08]**** | 29.0 | 3.9 [3.77-4.01]**** | 35.1 | 4.7 [4.56-4.85]**** |
| HT | 48,110 (42.1)) | 12,919 (56.8) | 20.4 | 2.7 [2.63-2.84]**** | 27.0 | 3.6 [3.51-3.73]**** | 34.0 | 4.6 [4.42-4.70]**** |
| CVD | 5,550 (4.9) | 1,681 (7.4) | 18.0 | 2.4 [2.09-2.76]**** | 26.7 | 3.6 [3.29-3.89]**** | 35.9 | 4.8 [4.57-5.07]**** |
| CKD | 5,152 (4.5) | 2,031 (8.9) | 21.8 | 2.9 [2.54-3.35]**** | 35.5 | 4.8 [4.41-5.13]**** | 45.1 | 6.1 [5.79-6.31]**** |
| Smoker | 23,490 (20.6) | 3,451 (15.2) | 6.7 | 0.9 [0.83-0.96]** | 15.6 | 2.1 [1.97-2.21]**** | 31.4 | 4.2 [4.02-4.40]**** |
| Asthma | 4,824 (4.2) | 469 (2.1) | 4.1 | 0.6 [0.45-0.67]**** | 11.0 | 1.5 [1.26-1.70]**** | 20.2 | 2.7 [2.40-3.05]**** |
| COPD | 3,747 (3.3) | 1,375 (6.0) | 27.9 | 3.7 [3.31-4.21]**** | 34.8 | 4.7 [4.29-5.07]**** | 41.0 | 5.5 [5.18-5.81]**** |
| ImmunoS | 2,608 (2.3) | 732 (3.2) | 17.7 | 2.4 [2.01-2.78]**** | 24.9 | 3.3 [2.94-3.77]**** | 36.5 | 4.9 [4.52-5.29]**** |
| Other | 5,309 (4.6) | 1,503 (6.6) | 19.5 | 2.6 [2.39-2.87]**** | 27.8 | 3.7 [3.43-4.03]**** | 38.7 | 5.2 [4.86-5.51]**** |
| Individuals w/comorb[b] | 114,322 (100) | 22,756 (100) | 14.9 | 2.0 [1.94-2.05]**** | 23.9 | 3.2 [3.11-3.29]**** | 33.4 | 4.5 [4.35-4.61]**** |
| Females | | | | | | | | |
| Obesity | 42,611 (43.9) | 5,113 (36.6) | 5.4 | 1.7 [1.59-1.81]**** | 12.5 | 4.0 [3.75-4.20]**** | 24.4 | 7.7 [7.38-8.10]**** |
| Diabetes | 34,177 (35.2) | 7,775 (55.7) | 15.8 | 5.0 [4.74-5.32]**** | 22.0 | 7.0 [6.68-7.30]**** | 29.0 | 9.2 [8.80-9.59]**** |
| HT | 43,354 (44.6) | 9,170 (65.7) | 14.3 | 4.5 [4.29-4.77]**** | 21.0 | 6.6 [6.37-6.93]**** | 28.4 | 9.0 [8.63-9.39]**** |
| CVD | 4,174 (4.3) | 997 (7.1) | 9.8 | 3.1 [2.48-3.89]**** | 17.6 | 5.6 [1.94-2.05]**** | 30.7 | 9.7 [9.1-10.42]**** |
| CKD | 3,963 (4.1) | 1,372 (9.8) | 14.9 | 4.7 [3.86-5.74]**** | 28.3 | 9.0 [8.03-9.97]**** | 41.4 | 13.1 [12.4-13.9]**** |
| Smoker | 9,827 (10.1) | 643 (4.6) | 1.8 | 0.6 [0.47-0.71]**** | 6.4 | 2.0 [1.76-2.34]**** | 17.9 | 5.7 [5.13-6.24]**** |
| Asthma | 7,522 (7.7) | 564 (4.0) | 2.8 | 0.9 [0.74-1.09][ns] | 6.3 | 2.0 [1.69-2.36]**** | 17.4 | 5.5 [4.96-6.09]**** |
| COPD | 3,464 (3.6) | 1,051 (7.5) | 23.1 | 7.3 [6.27-8.49]**** | 26.6 | 8.4 [7.57-9.36]**** | 34.6 | 11.0 [10.2-11.8]**** |
| ImmunoS | 2,846 (2.9) | 603 (4.3) | 11.3 | 3.6 [2.95-4.33]**** | 18.5 | 5.9 [5.04-6.80]**** | 29.8 | 9.4 [8.6-10.33]**** |
| Other | 6,362 (6.6) | 1,093 (7.8) | 8.1 | 2.6 [2.23-2.92]**** | 16.7 | 5.3 [4.75-5.90]**** | 29.1 | 9.2 [8.55-9.93]**** |
| Individuals w/comorb[b] | 97,173 (100) | 13,959 (100) | 9.2 | 2.9 [2.81-3.05]**** | 17.9 | 5.7 [5.45-5.91]**** | 27.9 | 8.7 [8.31-9.02]**** |
| Total | | | | | | | | |
| Obesity | 86,729 (41.0) | 12,455 (33.9) | 8.2 | 1.5 [1.46-1.57]**** | 14.7 | 2.7 [2.62-2.81]**** | 26.8 | 5.0 [4.83-5.11]**** |
| Diabetes | 73,771 (34.9) | 19,128 (52.1) | 19.5 | 3.6 [3.49-3.72]**** | 25.7 | 4.8 [4.64-4.88]**** | 32.1 | 5.9 [5.79-6.09]**** |
| HT | 91,464 (43.2) | 22,089 (60.2) | 17.6 | 3.3 [3.16-3.35]**** | 24.1 | 4.5 [4.36-4.58]**** | 31.3 | 5.8 [5.65-5.93]**** |
| CVD | 9,724 (4.6) | 2,678 (7.3) | 14.5 | 2.7 [2.37-3.01]**** | 23.1 | 4.3 [3.97-4.58]**** | 33.6 | 6.2 [5.98-6.48]**** |
| CKD | 9,115 (4.3) | 3,403 (9.3) | 18.8 | 3.5 [3.10-3.90]**** | 32.5 | 6.0 [5.65-6.40]**** | 43.5 | 8.1 [7.78-8.33]**** |
| Smoker | 33,317 (15.8) | 4,094 (11.2) | 5.2 | 1.0 [0.9-1.04][ns] | 12.9 | 2.4 [2.27-2.52]**** | 27.5 | 5.1 [4.88-5.30]**** |
| Asthma | 12,346 (5.8) | 1,033 (2.8) | 3.4 | 0.6 [0.54-0.71]**** | 8.2 | 1.5 [1.35-1.69]**** | 18.4 | 3.4 [3.15-3.67]**** |
| COPD | 7,211 (3.4) | 2,426 (6.6) | 25.7 | 4.8 [4.33-5.23]**** | 30.9 | 5.7 [5.35-6.10]**** | 37.9 | 7.0 [6.71-7.32]**** |
| ImmunoS | 5,454 (2.6) | 1,335 (3.6) | 14.2 | 2.6 [2.32-2.97]**** | 21.6 | 4.0 [3.64-4.40]**** | 33.1 | 6.1 [5.76-6.49]**** |
| Other | 11,671 (5.5) | 2,596 (7.1) | 13.0 | 2.4 [2.23-2.61]**** | 22.0 | 4.1 [3.81-4.34]**** | 33.5 | 6.2 [5.91-6.51]**** |
| Individuals w/comorb[b] | 211,495 (100) | 36,715 (100) | 12.3 | 2.3 [2.23-2.33]**** | 21.1 | 3.9 [3.82-3.99]**** | 31.1 | 5.6 [5.51-5.78]**** |

[a] All RRs were calculated against individuals with no comorbidities, in the same gender, excluding individuals with unknown comorbidity status **** p<0.0001; ** p<0.01; ns, not significant; CI, confidence interval.

[b] Some patients had more than one comorbidity, thus the sum of comorbidities (n and %) is larger than the individuals with comorbidity (w/comorb). Percents were calculated using "individuals w/comorbidity" as 100%.

**Table S6.** Frequency, measured case fatality rate (mCFR) and relative risk of death ($RR_d$) of specific comorbidities by age group, in COVID-19 in Mexico until August/6/2020 (**first cutoff**).

| Comorbidity /Age group | Cases n (%) | Deaths n (%) | As a single comorbidity %m CFR | As a single comorbidity RR [95% CI] | In combination with 1 other comorbidity %m CFR | In combination with 1 other comorbidity RR [95% CI] | In combination with ≥2 other comorbidities %m CFR | In combination with ≥2 other comorbidities RR [95% CI] |
|---|---|---|---|---|---|---|---|---|
| 0-19 yo | | | | | | | | |
| Obesity | 839 (44.3) | 18 (18.0) | 1.8 | 2.3 [1.3-4.17][**] | 2.9 | 3.7 [1.43-9.5][**] | 4.4 | 5.6 [1.52-18.9][**] |
| Diabetes | 151 (8.0) | 8 (8.0) | 1.2 | 1.6 [0.27-8.5][ns] | 9.1 | 11.6 [4.5-27.7][****] | 12.0 | 15.3 [5.3-39.2][****] |
| HT | 124 (6.6) | 9 (9.0) | 4.4 | 5.6 [1.5-18.9][**] | 9.8 | 12.5 [4.9-29.5][****] | 8.1 | 10.4 [3.5-27.8][****] |
| CVD | 154 (8.1) | 8 (8.0) | 3.5 | 4.4 [1.5-12.6][**] | 5.9 | 7.5 [2.6-20.7][****] | 12.5 | 16.0 [4.4-47.1][****] |
| CKD | 93 (4.9) | 11 (11.0) | 4.4 | 5.6 [1.5-18.9][**] | 26.9 | 34.4 [17.1-61.3][****] | 4.8 | 6.1 [1.07-29.4][*] |
| Smoker | 266 (14.0) | 2 (2.0) | - | - | 4.3 | 5.4 [1.5-18.5][**] | - | - |
| Asthma | 567 (29.9) | 2 (2.0) | 0.4 | 0.55 [0.15-2.0][ns] | - | - | - | - |
| COPD[b] | 16 (0.8) | 0 (0.0) | - | - | - | - | - | - |
| ImmunoS | 311 (16.4) | 28 (28.0) | 5.6 | 7.1 [3.8-13.1][****] | 13.3 | 17.0 [9.8-28.4][****] | 15.2 | 19.4 [8.3-40.7][****] |
| Other | 449 (23.7) | 54 (54.0) | 12.5 | 15.9 [11.0-22.8][****] | 11.2 | 14.3 [8.5-23.5][****] | 11.8 | 15.0 [5.9-34.9][****] |
| Individuals w/comorb[c] | 1,894 (100) | 100 (100) | 3.3 | 4.2 [3.1-5.65][****] | 7.8 | 10.0 [6.6-15.05][****] | 9.5 | 12.1 [5.8-24.0][****] |
| 20-39 yo | | | | | | | | |
| Obesity | 27,478 (53.0) | 1,032 (55.4) | 3.0 | 3.2 [2.9-3.58][****] | 4.2 | 4.6 [4.0-5.2][****] | 9.5 | 10.4 [9.0-12.1][****] |
| Diabetes | 5,954 (11.5) | 541 (29.1) | 6.4 | 7.0 [6.0-8.25][****] | 8.8 | 9.6 [8.23-11.2][****] | 14.5 | 16.0 [13.8-18.4][****] |
| HT | 8,213 (15.8) | 564 (30.3) | 2.9 | 3.2 [2.6-3.94][****] | 7.2 | 8.0 [6.9-9.1][****] | 12.9 | 14.1 [12.4-16.2][****] |
| CVD | 1,089 (2.1) | 67 (3.6) | 2.6 | 2.9 [1.61-5.1][***] | 3.6 | 4.0 [2.3-6.9][****] | 13.0 | 14.3 [10.7-18.8][****] |
| CKD | 1,579 (3.0) | 275 (14.8) | 9.8 | 10.8 [8.3-14.0][****] | 19.0 | 20.9 [17.5-24.9][****] | 24.0 | 26.3 [22.1-31.1][****] |
| Smoker | 14,125 (27.2) | 228 (12.2) | 0.6 | 0.7 [0.5-0.91][**] | 2.5 | 2.7 [2.2-3.4][****] | 6.9 | 7.6 [6.0-9.5][****] |
| Asthma | 4,994 (9.6) | 97 (5.2) | 0.9 | 1.0 [0.66-1.4][ns] | 2.8 | 3.1 [2.2-4.2][****] | 5.9 | 6.6 [4.55-9.1][****] |
| COPD[b] | 395 (0.8) | 33 (1.8) | 3.2 | 3.5 [1.37-8.7][**] | 4.7 | 5.2 [2.4-10.8][****] | 16.2 | 17.8 [12.1-25.5][****] |
| ImmunoS | 1,130 (2.2) | 128 (6.9) | 6.2 | 6.8 [4.70-9.8][****] | 9.9 | 10.9 [7.8-15.05][****] | 18.7 | 20.6 [16.4-25.5][****] |
| Other | 2,942 (5.7) | 159 (8.5) | 2.9 | 3.2 [2.4-4.25][****] | 6.4 | 7.0 [5.4-9.2][****] | 12.8 | 14.0 [10.9-17.8][****] |
| Individuals w/comorb[c] | 51,869 (100) | 1,862 (100) | 2.6 | 2.8 [2.6-3.09][****] | 5.3 | 5.9 [5.3-6.5][****] | 11.9 | 13.1 [11.6-14.7][****] |
| 40-59 yo | | | | | | | | |
| Obesity | 41,075 (44.2) | 5,383 (42.8) | 9.5 | 1.6 [1.54-1.71][****] | 12.7 | 2.2 [2.1-2.3][****] | 20.7 | 3.5 [3.4-3.7][****] |
| Diabetes | 34,772 (37.4) | 6,574 (52.3) | 15.1 | 2.8 [2.45-2.71][****] | 17.9 | 3.1 [2.9-3.2][****] | 24.6 | 4.2 [4.0-4.4][****] |
| HT | 40,419 (43.5) | 6,488 (51.6) | 9.3 | 1.6 [1.49-1.68][****] | 16.3 | 2.8 [2.7-2.9][****] | 23.8 | 4.1 [3.9-4.2][****] |
| CVD | 3,099 (3.3) | 546 (4.3) | 8.2 | 1.4 [1.06-1.83][*] | 12.4 | 2.1 [1.8-2.5][****] | 23.6 | 4.0 [3.7-4.4][****] |
| CKD | 3,438 (3.7) | 1,226 (9.8) | 20.6 | 3.5 [2.92-4.19][****] | 29.3 | 5.0 [4.5-5.6][****] | 41.2 | 7.1 [6.65-7.45][****] |
| Smoker | 12,250 (13.2) | 1,353 (10.8) | 5.5 | 0.9 [0.84-1.06][ns] | 11.8 | 2.0 [1.85-2.2][****] | 19.8 | 3.4 [3.1-3.65][****] |
| Asthma | 4,907 (5.3) | 438 (3.5) | 4.3 | 0.7 [0.59-0.90][**] | 8.1 | 1.4 [1.2-1.6][***] | 15.9 | 2.7 [2.4-3.1][****] |
| COPD | 1,911 (2.6) | 425 (3.4) | 17.1 | 2.9 [2.34-3.61][****] | 15.8 | 2.7 [2.2-3.3][****] | 28.2 | 4.8 [4.3-5.3][****] |
| ImmunoS | 2,219 (2.4) | 462 (3.7) | 13.9 | 2.4 [1.95-2.89][****] | 17.0 | 2.9 [2.4-3.45][****] | 27.8 | 4.8 [4.3-5.3][****] |
| Other | 4,707 (5.1) | 817 (6.5) | 11.4 | 1.9 [1.70-2.21][****] | 15.6 | 2.7 [2.35-3.0][****] | 26.1 | 4.5 [4.1-4.9][****] |
| Individuals w/comorb[b] | 92,928 (100) | 12,569 (100) | 10.3 | 1.8 [1.70-1.83][****] | 15.3 | 2.6 [2.5-2.7][****] | 23.3 | 4.0 [3.8-4.1][****] |
| 60-79 yo | | | | | | | | |
| Obesity | 15,869 (28.6) | 5,381 (29.5) | 28.0 | 1.16 [1.10-1.23][****] | 32.3 | 1.34 [1.28-1.40][****] | 37.8 | 1.57 [1.52-1.63][****] |
| Diabetes | 29,225 (52.7) | 10,330 (56.7) | 30.2 | 1.26 [1.20-1.31][****] | 34.4 | 1.43 [1.38-1.48][****] | 39.7 | 1.65 [1.60-1.71][****] |
| HT | 36,287 (65.4) | 12,146 (66.6) | 27.6 | 1.15 [1.10-1.19][****] | 33.1 | 1.38 [1.33-1.42][****] | 39.1 | 1.63 [1.57-1.68][****] |
| CVD | 4,202 (7.6) | 1,530 (8.4) | 26.8 | 1.11 [0.95-1.30][ns] | 32.0 | 1.33 [1.21-1.46][****] | 39.6 | 1.65 [1.56-1.73][****] |
| CKD | 3,438 (6.2) | 1,592 (8.7) | 35.2 | 1.47 [1.21-1.74][***] | 43.9 | 1.83 [1.67-1.99][****] | 47.8 | 2.00 [1.90-2.10][****] |
| Smoker | 5,724 (10.3) | 2,065 (11.3) | 29.7 | 1.24 [1.14-1.34][****] | 33.8 | 1.41 [1.31-1.51][****] | 40.9 | 1.70 [1.61-1.79][****] |
| Asthma | 1,656 (3.0) | 428 (2.3) | 22.0 | 0.92 [0.75-1.11][ns] | 23.8 | 0.99 [0.83-1.16][ns] | 28.5 | 1.19 [1.06-1.32][**] |
| COPD | 3,591 (6.5) | 1,395 (7.7) | 29.8 | 1.24 [1.08-1.41][**] | 36.6 | 1.52 [1.40-1.65][****] | 42.3 | 1.76 [1.66-1.86][****] |
| ImmunoS | 1,550 (2.8) | 593 (3.3) | 28.5 | 1.18 [0.97-1.43][ns] | 35.1 | 1.46 [1.28-1.66][****] | 42.7 | 1.78 [1.64-1.92][****] |
| Other | 2,919 (5.3) | 1,213 (6.7) | 37.4 | 1.56 [1.40-1.72][****] | 40.7 | 1.69 [1.56-1.84][****] | 43.9 | 1.83 [1.71-1.94][****] |
| Individuals w/comorb[b] | 55,466 (100) | 18,234 (100) | 28.8 | 1.20 [1.16-1.23][****] | 33.8 | 1.40 [1.36-1.45][****] | 39.2 | 1.63 [1.58-1.68][****] |
| ≥80 yo | | | | | | | | |
| Obesity | 1,468 (16.7) | 641 (16.2) | 41.2 | 1.07 [0.91-1.25][ns] | 43.4 | 1.13 [1.01-1.26][*] | 44.5 | 1.16 [1.06-1.27][**] |
| Diabetes | 3,669 (41.8) | 1,675 (42.4) | 44.0 | 1.15 [1.04-1.26][**] | 45.2 | 1.18 [1.10-1.26][****] | 46.9 | 1.21 [1.14-1.31][****] |
| HT | 6,421 (73.2) | 2,882 (73.0) | 43.4 | 1.13 [1.06-1.21][***] | 44.8 | 1.17 [1.10-1.24][****] | 46.9 | 1.22 [1.14-1.31][****] |
| CVD | 1,180 (13.5) | 527 (13.3) | 48.1 | 1.25 [1.03-1.49][*] | 43.0 | 1.12 [0.98-1.27][ns] | 44.9 | 1.17 [1.06-1.28][**] |
| CKD | 567 (6.5) | 299 (7.6) | 44.2 | 1.15 [0.79-1.54][ns] | 56.3 | 1.47 [1.24-1.69][****] | 52.4 | 1.37 [1.23-1.51][****] |
| Smoker | 952 (10.8) | 446 (11.3) | 44.4 | 1.16 [0.97-1.35][ns] | 47.1 | 1.23 [1.07-1.39][**] | 47.7 | 1.24 [1.12-1.38][****] |

| | | | | | | | | |
|---|---|---|---|---|---|---|---|---|
| Asthma | 222 (2.5) | 68 (1.7) | 22.6 | 0.59 [0.30-1.04]$^{ns}$ | 27.7 | 0.72 [0.48-1.04]$^{ns}$ | 34.1 | 0.89 [0.69-1.12]$^{ns}$ |
| COPD | 1,298 (14.8) | 573 (14.5) | 45.2 | 1.18 [1.01-1.36]$^{*}$ | 45.2 | 1.18 [1.05-1.31]$^{**}$ | 43.1 | 1.13 [1.02-1.24]$^{*}$ |
| ImmunoS | 244 (2.8) | 124 (3.1) | 50.0 | 1.30 [0.93-1.68]$^{ns}$ | 50.0 | 1.30 [0.99-1.62]$^{ns}$ | 51.5 | 1.34 [1.12-1.57]$^{**}$ |
| Other | 654 (7.5) | 353 (8.9) | 55.8 | 1.45 [1.21-1.70]$^{***}$ | 54.9 | 1.43 [1.24-1.63]$^{****}$ | 52.9 | 1.38 [1.23-1.53]$^{****}$ |
| Individuals w/comorb[b] | 8,776 (100) | 3,950 (100) | 44.0 | 1.15 [1.08-1.21]$^{****}$ | 45.3 | 1.18 [1.11-1.25]$^{****}$ | 46.6 | 1.22 [1.14-1.30]$^{****}$ |

[a] All RRs were calculated against individuals with no comorbidities, in the same age-group, excluding individuals with unknown comorbidity status, **** p<0.0001; *** p<0.001; ** p<0.01; * p<0.05; ns, not significant; CI, confidence interval. [b] Some patients had more than one comorbidity, thus the sum of comorbidities (n and %) is larger than the individuals with comorbidity (w/comorb). Percents were calculated using "individuals w/comorbidity" as 100%.

**Table S7.** Frequency of the 23 most common comorbidity combinations by age group, in Mexico until Sept/12/2020 (second cutoff).

| Comorbidity | 20-39 yo | | 40-59 yo | | 60-79 yo | | ≥80 yo | |
|---|---|---|---|---|---|---|---|---|
| | Cases (%) | Deaths (%) | Cases (%) | Deaths (%) | Cases(%) | Deaths (%) | Cases (%) | Deaths (%) |
| No comorb. | 173,011(69.8) | 1,420(36.5) | 129,754(50.0) | 7,141(29.6) | 33,694(30.1) | 8,068(23.6) | 4,580(26.80) | 1,786(23.5) |
| DM-HT | 881 (0.4) | 85 (2.18) | 10,769 (4.2) | 2,022 (8.4) | 13,436 (12.0) | 4,602(13.4) | 1,992(11.7) | 928(12.21) |
| DM-obesity | 1,444 (0.6) | 109 (2.80) | 4,822 (1.86) | 709 (2.94) | 1,847 (1.65) | 639 (1.87) | 110 (0.64) | 57 (0.75) |
| Obesity-HT | 2,680 (1.1) | 118 (3.03) | 8,831 (3.40) | 1,082 (4.5) | 4,167 (3.72) | 1,354(3.95) | 440 (2.57) | 194 (2.55) |
| DM-CKD | 70 (0.03) | 15 (0.39) | 272 (0.10) | 110 (0.46) | 325 (0.29) | 171 (0.50) | 32 (0.19) | 14 (0.18) |
| HT-CKD | 450 (0.18) | 106 (2.72) | 555 (0.21) | 175 (0.73) | 465 (0.42) | 191 (0.56) | 143 (0.84) | 80 (1.05) |
| Obesity-CKD | 98 (0.04) | 7 (0.18) | 111 (0.04) | 22 (0.09) | 48 (0.04) | 19 (0.06) | 2 (0.01) | 1 (0.01) |
| CKD-ImmunoS | 70 (0.03) | 15 (0.39) | 41 (0.02) | 9 (0.04) | 7 (0.01) | 2 (0.01) | 3 (0.02) | 1 (0.01) |
| DM-smoker | 267 (0.11) | 18 (0.46) | 1,089 (0.42) | 160 (0.66) | 621 (0.55) | 221 (0.65) | 72 (0.42) | 43 (0.57) |
| HT-smoker | 338 (0.14) | 14 (0.36) | 1,047 (0.40) | 106 (0.44) | 829 (0.74) | 256 (0.75) | 191(1.12) | 86 (1.13) |
| Ob-smoker | 4,208 (1.70) | 85 (2.18) | 2,929 (1.1) | 298 (1.24) | 482 (0.43) | 149 (0.43) | 30 (0.18) | 18 (0.24) |
| DM-COPD | 23 (0.01) | 2 (0.05) | 160 (0.06) | 32 (0.13) | 346 (0.31) | 139 (0.41) | 91 (0.53) | 42 (0.55) |
| HT-COPD | 25 (0.01) | 2 (0.05) | 212 (0.08) | 38 (0.16) | 564 (0.50) | 216 (0.63) | 285 (1.67) | 142 (1.87) |
| Ob-COPD | 57 (0.02) | 2 (0.05) | 180 (0.07) | 29 (0.12) | 161 (0.14) | 50 (0.15) | 35 (0.20) | 13 (0.17) |
| Ob-asthma | 1,127 (0.45) | 31 (0.80) | 922 (0.36) | 68 (0.28) | 131 (0.12) | 30 (0.09) | 9 (0.05) | 0 (0.00) |
| HT-CVD | 83 (0.03) | 4 (0.10) | 475 (0.18) | 50 (0.21) | 839 (0.75) | 269 (0.79) | 355 (2.08) | 151 (1.99) |
| Ob-DM-HT | 740 (0.30) | 80 (2.05) | 5,998 (2.31) | 1,165 (4.8) | 4,709 (4.2) | 1,696 (5.0) | 377 (2.21) | 184 (2.42) |
| Ob-DM-HT-CKD | 28 (0.01) | 8 (0.21) | 308 (0.12) | 128 (0.53) | 388 (0.35) | 200 (0.58) | 28 (0.16) | 11 (0.14) |
| Ob-DM-HT-smkr | 91 (0.04) | 4 (0.10) | 548 (0.21) | 101 (0.42) | 395 (0.35) | 170 (0.50) | 37 (0.22) | 23 (0.30) |
| Ob-DM-HT-CVD | 21 (0.01) | 1 (0.03) | 284 (0.11) | 71 (0.29) | 492 (0.44) | 187 (0.55) | 75 (0.44) | 31 (0.41) |
| Ob-DM-HT-COPD | 7 (0.003) | 3 (0.08) | 127 (0.05) | 44 (0.18) | 281 (0.25) | 117 (0.34) | 49 (0.29) | 21 (0.28) |
| DM-HT-CKD | 130 (0.05) | 48 (1.23) | 1,223 (0.47) | 523 (2.17) | 1,565 (1.40) | 765 (2.23) | 164 (0.96) | 91 (1.20) |
| DM-HT-CKD-smkr | 7 (0.003) | 3 (0.08) | 71 (0.03) | 37 (0.15) | 103 (0.09) | 46 (0.13) | 8 (0.05) | 7 (0.09) |
| DM-HT-CVD | 25 (0.01) | 3 (0.08) | 397 (0.15) | 78 (0.32) | 881 (0.79) | 300 (0.88) | 226 (1.32) | 108 (1.4) |

[a]All RRs were calculated against the population with no comorbidities in the age-group. Data are for the second cutoff date (Sept/12/2020), excluding individuals with unknown comorbidity status, as in Tables 1 and 2. **** p<0.0001; *** p<0.001; ** p<0.01; * p<0.05; ns, not significant; CI, confidence interval. Disease abbreviations are as Table 1.

**Table S8.** Years of life lost to premature COVID-19 mortality (YLL) in Mexico until end of study (Nov/2/2020), subdivided by gender and presence/absence of comorbidity.

| | YLL [a] (%) | | | Number of people with YLL (%) | | | EXCESS YLL FROM COMORB. (% of total YLL) [b] |
|---|---|---|---|---|---|---|---|
| | No Comorbidity | With Comorbidity | Total YLL | No comorbidity | With Comorbidity | Total Individuals | |
| Females | 75,033 (25.3) | 221,209 (74.7) | 296,242 (100) | 4,555 (22.0) | 16,170 (78.0) | 20,725 (100) | 146,176 (16.6) |
| Males | 196,434 (33.7) | 386,735 (66.3) | 583,169 (100) | 11,955 (31.0) | 26,649 (69.0) | 38,604 (100) | 190,301 (21.6) |
| Total | 271,467 (30.9) | 607,944 (69.1) | 879,411 (100) | 16,510 (27.8) | 42,819 (72.2) | 59,329 (100) | 336,477 (38.3) |

[a] For each deceased individual under age 70, YLL were calculated with the formula: YLL=(70 years)-(age of deceased individual).

[b] Excess YLL from the synergy between COVID-19 and the studied comorbidities in Mexico, were estimated by calculating the difference between YLL with and without comorbitity, with the formula: Excess YLL=(YLL with comorbidity)-(YLL without comorbidity). Percent in the last column were calculated using the total YLL (879,411) as 100%.

**Table S9.** Age stratified comparison of diabetes and obesity prevalence in Mexico *vs* the USA, as reported by health authorities.

| Diabetes | | | |
|---|---|---|---|
| Mexico | | USA | |
| Age group | % prevalence | Age group | % prevalence |
| 20-39 | 1.8 | 18-44 | 3.0 |
| 40-59 | 12.8 | 45-64 | 13.8 |
| >60 | 25.1 | ≥65 | 21.4 |
| Total | 10.3 | Total | 9.9 |

| Obesity | % prevalence | | | |
|---|---|---|---|---|
| | Mexico | | USA | |
| Age group | males | females | males | females |
| 5-11 | 20.1 | 15.0 | 18.5 | |
| 12-19 | 15.1 | 14.1 | | |
| 20-29 | 24.1 | 26.2 | 34.8 | 36.5 |
| 30-39 | 34.0 | 39.0 | | |
| 40-49 | 37.4 | 50.0 | 40.8 | 44.7 |
| 50-59 | 32.8 | 47.3 | | |
| 60-69 | 31.1 | 48.4 | 38.5 | 43.1 |
| 70-79 | 23.2 | 33.1 | | |
| >80 | 12.5 | 17.6 | | |

### Supplementary Figures

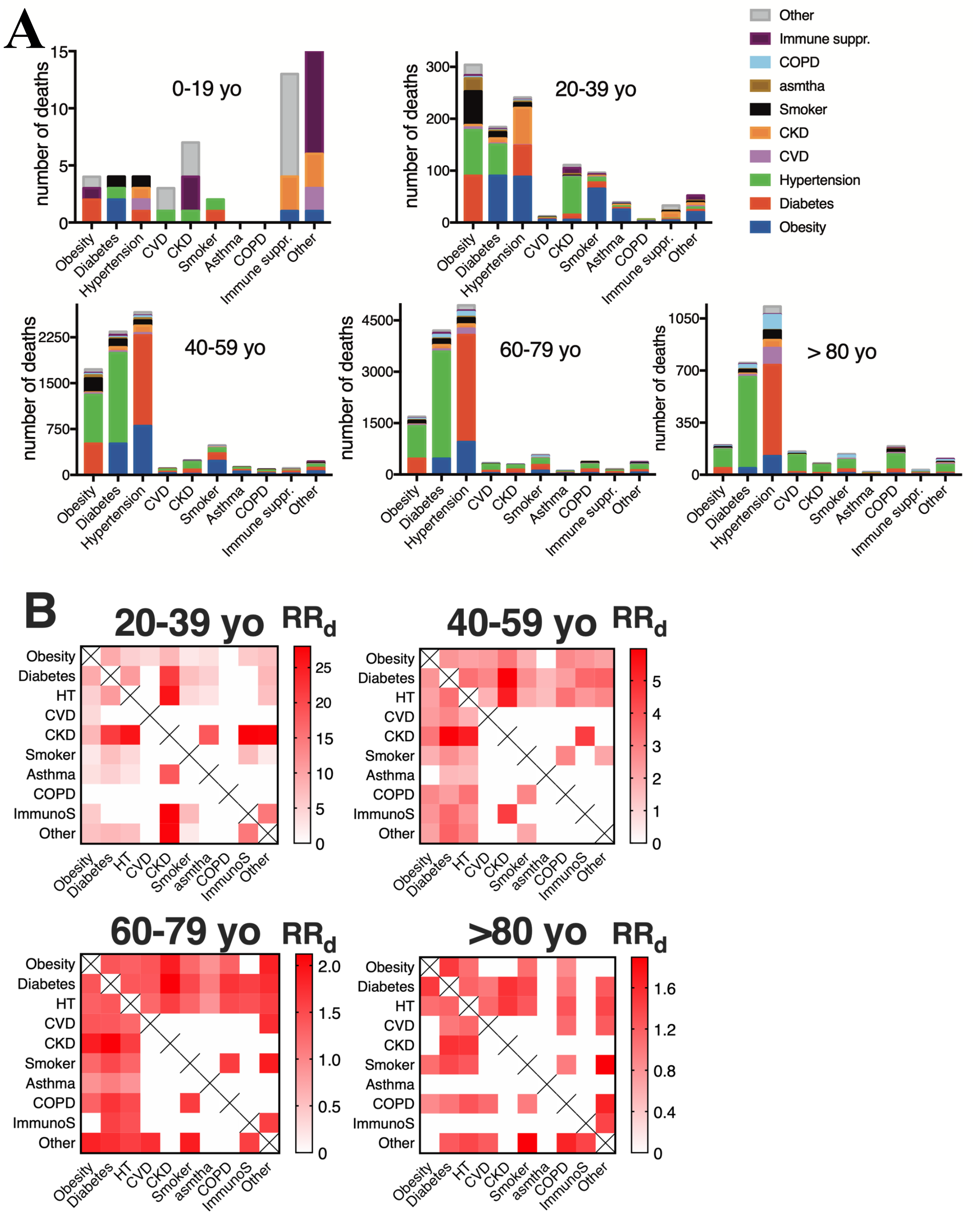


**Figure S1 Combinations of 2 comorbidities in COVID-19 deaths. (A)** Combinations sorted by age, until Aug/6/2020 (first cutoff). This is a bar representation of a symmetric matrix (same diseases in columns and rows) to allow visualization of frequent combinations. **(B)** the RRd of the 25-27 most common disease combinations are represented. Infrequent combinations (present in <0.2% of deaths in the age group) were excluded (blank squares) due to large 95% CI. A different RRd scale was used for each age-group, because the RRd at older ages is smaller.

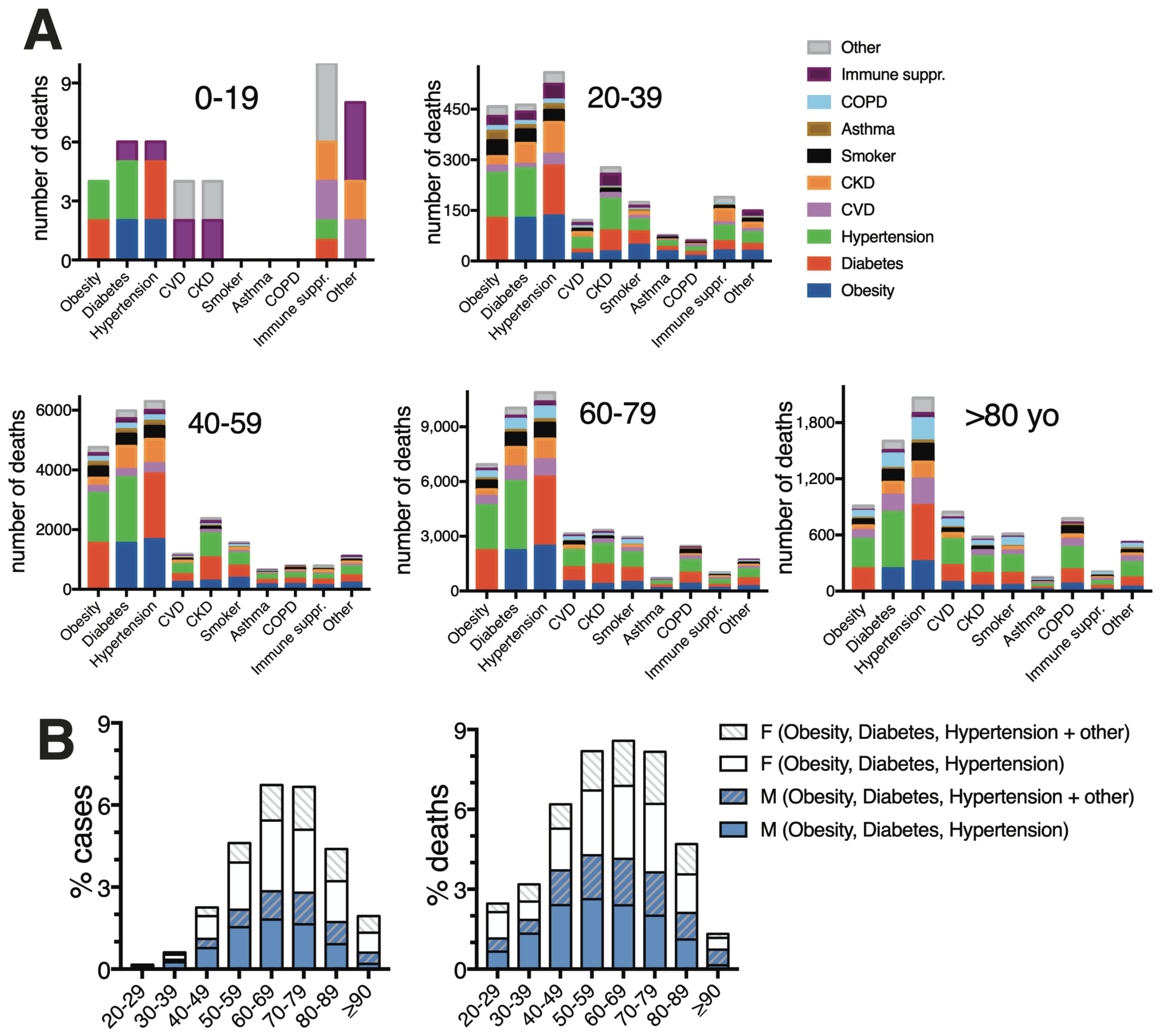


**Figure S2 Combinations of 3 or more comorbidities in COVID-19 deaths. (A)** Combinations of pre-existing diseases, in individuals that died from COVID-19 and had **3 or more** comorbidities, depicted by age group, until Aug/6/2020 (first cutoff). This is a bar representation of a symmetric matrix (same diseases in columns and rows). **(B)** Percent of total COVID-19 cases and deaths that had the triad of obesity/diabetes/hypertension, with gender distribution ± other disease(s).

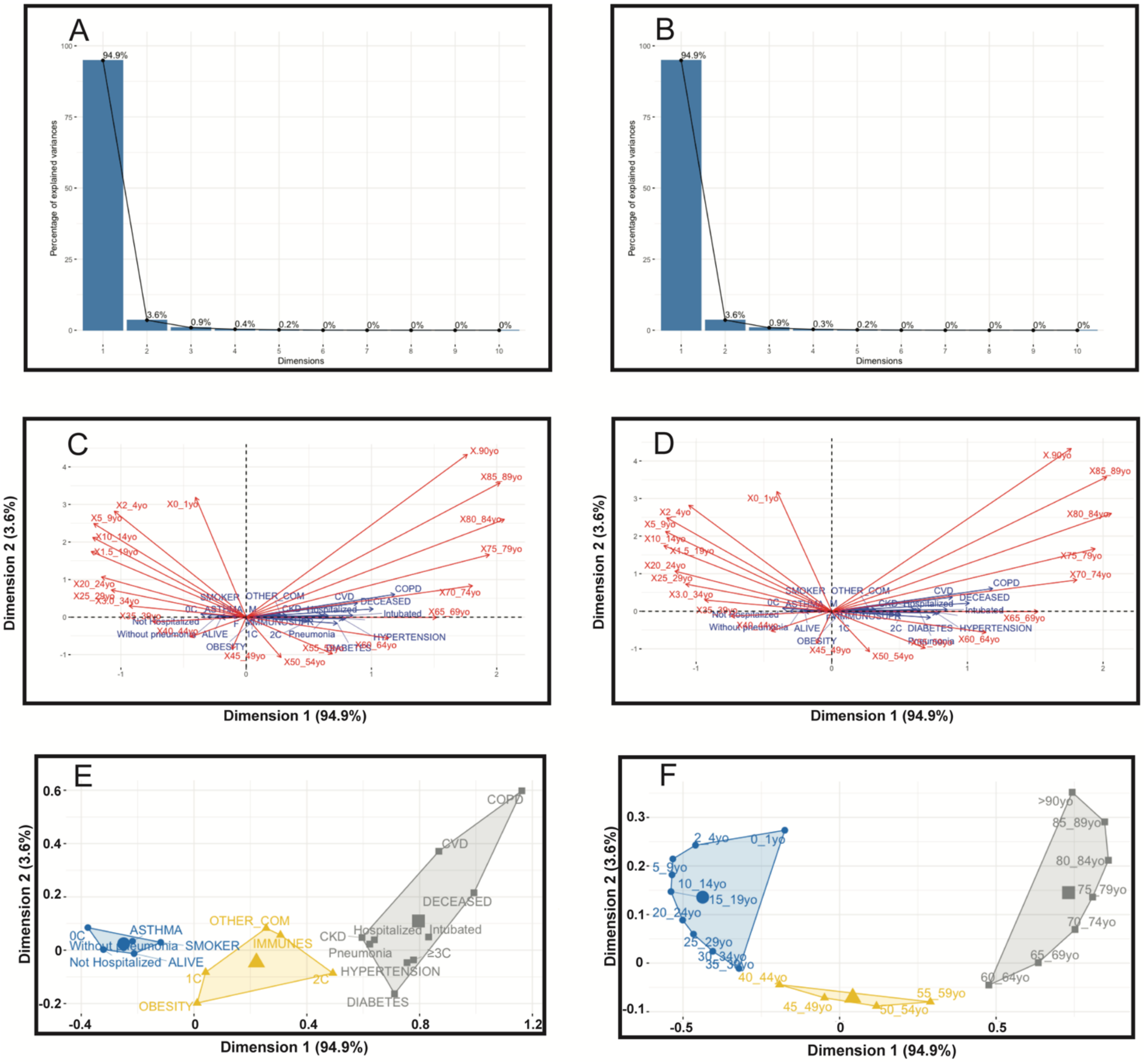


**Figure S3 Correspondence Analyses and Hierarchical Clustering of Principal Components (HCPC) of the Mexican COVID-19 datasets**. Datasets were cutoff at: Aug/6 (**A,C**) and Sept/12/2020 (**B,D,E,F**). (**A-B**) The first two dimensions obtained by CA explain 98.5% of the total variation (94.9% for the first dimension and 3.6% for the second). Differences between cutoffs are observed until the 4th dimension. (**C-D**) CA asymmetric biplots created by representing age-groups (columns) in row space (comorbidities and outcomes). (**E**) HCPC found three clusters relating different comorbidities and outcomes. Favorable and adverse outcomes were located at opposite ends of the x-axis (largest dimension). Disease abbreviations are as in text. In (**F**) the same data was transposed to show the clustering of ages relative to the comorbidities and outcomes displayed in (**E**). Clusters reflect how the different comorbidities and outcomes are represented in different age groups. CA clusters were indistinguishable between cutoff dates.
